# One blood methylation profile stratifies ten-year risk across 21 common diseases

**DOI:** 10.64898/2026.11.18.26368164

**Authors:** Kirsten B. Seale, Sayf Hassouneh, Laura Balagué-Dobón, Max Melnikas, Andrea Aparicio, Tao Guo, Kerry L. Ivey, Jessica A. Lasky-Su, Anne Richmond, Riccardo E. Marioni, Mike Mallin, Varun B. Dwaraka, Natàlia Carreras-Gallo, Ryan Smith

## Abstract

Blood DNA methylation offers a dynamic readout of inherited and acquired influences on health, creating an opportunity for risk prediction beyond polygenic scores, which are fixed at conception and have yet to demonstrate broad clinical utility. As such, we aimed to develop methylation risk scores for 21 common diseases in 12,785 adults from Generation Scotland and the Mass General Brigham Biobank, combining technically reproducible probes, null-calibrated feature selection and ten-year survival models, with evaluation in a single holdout that was not used for feature selection, model fitting or tuning. The methylation risk scores for eight diseases reached a holdout C-index of at least 0.80: chronic kidney disease, peripheral vascular disease, chronic obstructive pulmonary disease, heart failure and type 2 diabetes with at least 50 events each, and interstitial lung disease, dementia and fatty liver disease with fewer. For the five scores with at least 50 events, adjusted hazard ratios per standard deviation ranged from 2.6 to 4.1, with the highest risk tertile containing 73 to 90% of the ten-year events. Panel CpGs were enriched for cytokine signalling, innate immune sensing and sterol metabolism. Against reported values, the eight top-tier methylation risk scores exceed published polygenic and methylation scores alone, and exceed published polygenic and methylation scores that include clinical covariates for seven of the eight diseases (the exception is type 2 diabetes, where our AUC of 0.865 without clinical risk factors is close to a methylation-plus-risk-factor model at 0.872, and our C-index of 0.809 is lower than the C-index for a polygenic-plus-risk-factor at 0.85). These results indicate that a single blood methylation profile can rank ten-year risk for several common diseases at a level that compares favourably with published genetic and methylation scores.

## INTRODUCTION

The accurate identification of individuals at risk for disease is central to prevention. Molecular risk markers are usually studied in one of two designs: i) prevalent (cross-sectional) analyses compare people who already have a disease with people who do not, challenging our ability to separate markers of risk from consequences of the disease or its treatment, or ii) incident analyses, which follow people who are initially unaffected until some develop the disease, predicting future disease risk. The second is what is valuable in answering the clinical question, yet most risk scores have been evaluated with the first.

Most common chronic diseases are polygenic, and genome-wide association studies have identified thousands of variants that each confer a small shift in risk.^1^ Combined into a polygenic risk score (PRS), these variants stratify populations for coronary artery disease (CAD), type 2 diabetes (T2DM), breast cancer and other conditions, and for a few per cent of individuals a high PRS carries risk comparable to a monogenic mutation.^2,3^ Several limitations follow from how PRS are built. PRS capture inherited susceptibility, which is fixed at conception. This is an advantage for early risk assessment, but it also means that a PRS alone cannot register the lifestyle, environmental, metabolic and treatment exposures that determine when, or whether, inherited susceptibility becomes disease, nor change after an intervention. A systematic review of 591 studies found strong evidence of clinical validity for PRS in 22 but no study that demonstrated clinical utility, that is, improved outcomes from acting on the score.^4,5^ Many PRS have also been developed and evaluated in prevalent case-control studies, so their discrimination reflects lifetime susceptibility rather than risk over a defined horizon,^6^ which limits their value for modifiable, environmentally influenced diseases. Their discrimination in the general population is modest: standalone PRS for most common diseases reach an area under the curve (AUC) of 0.58–0.71 in UK Biobank, and most of the predictive value of a clinical PRS model comes from the age, sex and clinical covariates it is combined with.^6,7^ As with most risk scores, they transfer less well across ancestries, as discovery cohorts are predominantly European.^8^ A complementary, dynamic measure of acquired risk could therefore add information that inherited risk cannot.

DNA methylation measured in blood records both inherited and acquired influences on health, and unlike genotype, it changes over time. Methylation at specific CpG sites tracks smoking exposure and its cessation,^9^ body mass, alcohol use and other behaviours,^10^ circulating protein levels and cell composition,^11,12^ and methylation-based measures of biological ageing respond to randomised interventions such as caloric restriction.^13^ Predictors built on methylation therefore report on an individual’s current biological state rather than on a fixed liability. This is what gives a methylation risk score (MRS) its potential for clinical utility: the same assay that stratifies risk can be repeated to show whether risk has changed, and the CpGs and pathways a score relies on point to the biology that drives the risk. Metabolomic and proteomic scores are also dynamic and predict many incident diseases,^14,15^ however, the advantage of methylation is practical, whereby a single array on a small blood sample yields many predictors,^12,16^ which have already been used to estimate exposures and biomarkers that are costly or impractical to measure directly,^10,12^ to classify rare genetic disease,^17^ and to predict mortality^18^ and incident disease.^19,20^

Methylation-based predictors have developed along two lines. They were first trained on outcomes, such as chronological age or mortality, and then asked whether the score was associated with disease. Epigenetic clocks of this kind are associated with many conditions, but because they capture what diseases share rather than what distinguishes them, adding little disease-specific prediction. For example, in a test of 14 clocks against 174 incident diseases in Generation Scotland (GS), adding a clock to age, sex and lifestyle improved the 10-year AUC by more than 0.01 for only 32 of 176 significant clock-disease associations.^21^ The second line trained scores directly on a single disease. These disease-specific MRS have shown that blood methylation carries a usable signal. An example is a methylation score that raised the 10-year AUC for T2DM from 0.839 to 0.872 over clinical risk factors in GS and replicated in KORA;^22^ CpG panels improved lung cancer prediction over smoking status, and improved breast cancer prediction over self-reported risk factors and a PRS alone.^23,24^ Methylation scores for cardiovascular disease (CVD) have been built in several cohorts, including most recently, in the Million Veteran Program (MVP).^19,20,25^ The one study to build methylation scores for many phenotypes at once did so cross-sectionally in 831 patients and found that they outperformed PRS for most medication, laboratory and diagnosis codes.^26^

Those studies also show where the field has been limited. Each disease-specific MRS was built for one disease in one development cohort, with its own feature-selection and evaluation choices, so performance cannot be compared across diseases, and few studies have tested transfer to a second cohort.^19,22^ Sample sizes of a few hundred to a few thousand have meant that feature selection and evaluation often used the same participants, which inflates apparent accuracy,^27^ and that held-out evaluation with enough events to estimate discrimination with precision has rarely been possible. Most evaluations have used prevalent case-control status, as for PRS, rather than time-to-event outcomes, so the horizon over which a score predicts, which determines its clinical use, is usually unknown.

Modelling choices add a further limitation. Most methylation predictors consider marginal CpG effects or assume linear, additive effects between CpGs. Feature selection and modeling that capture interactions and non-linear effects could represent more complex relationships between CpGs and may improve discrimination.

Here we ask whether a single blood methylation profile can predict ten-year risk across many common chronic diseases when every score is built and tested in the same way. We developed methylation risk scores for 21 common diseases in 12,785 adults from two cohorts with different settings and outcome ascertainment, GS and the Mass General Brigham (MGB) Biobank, using one pipeline for probe selection, survival modelling and evaluation on a holdout set aside before any analysis. To our knowledge, this is the first uniform development and evaluation of methylation risk scores across this many diseases with incident ten-year outcomes. We report every score, including the weak ones, set the strongest against published methylation and polygenic scores, and examine the CpGs and pathways on which they rely.

## RESULTS

### Two cohorts provide 10-year incident outcomes for 21 diseases

We built separate methylation risk scores (MRS) for 21 diseases in 12,785 individuals from GS (n = 10,156) and MGB (n = 2,629) (Figure 1, Table 1).^28–30^ The cohorts respectively differed in age (mean 49.3 vs 57.2 years) and in follow-up of controls (median 15.1-15.2 vs 6.6-6.8 years). All results below are from a holdout of 1,920 individuals that was set aside before any analysis, with 28 to 168 incident events within 10 years per disease (Table 2).

**Figure 1.**
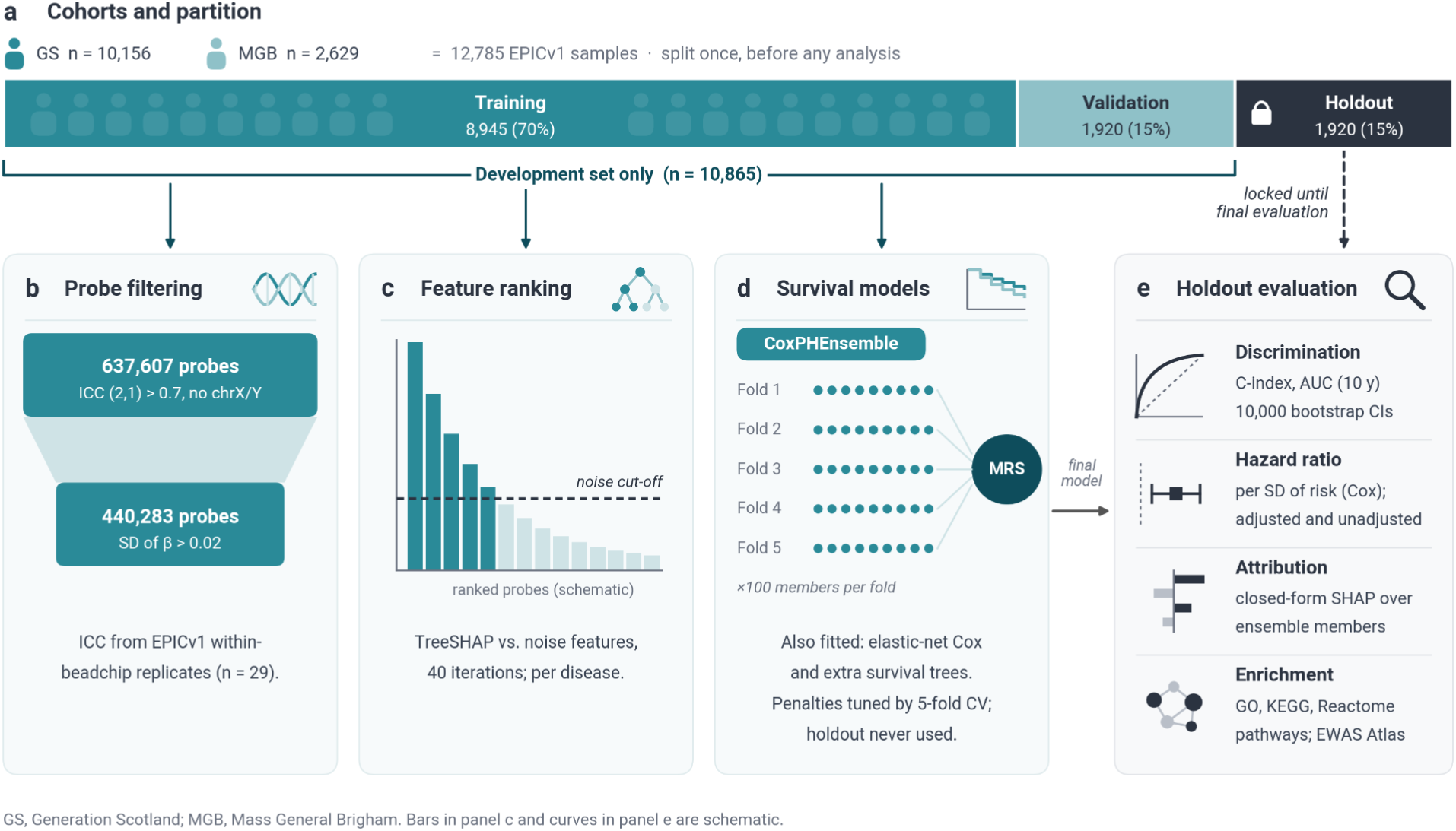
Study design for the development and evaluation of the methylation risk scores (MRS). (A) Whole-blood DNA methylation profiles from 12,785 individuals (Generation Scotland, GS, n = 10,156; Mass General Brigham, MGB, n = 2,629), measured on the Illumina EPICv1 array with one sample per individual. Samples were split once, before any analysis, into training (n = 8,945; 70%), validation (n = 1,920; 15%) and holdout (n = 1,920; 15%) partitions. Training and validation samples together formed the development set (n = 10,865). The holdout was set aside until final evaluation. (B) Probe filtering. Of the probes on the array, 637,607 passed an intraclass correlation coefficient threshold of ICC (2,1) > 0.7 (EPICv1 within-beadchip technical replicates, n = 29) and are not on chromosomes X or Y. A further filter on the standard deviation of beta values (> 0.02) left 440,283 probes. (C) Feature ranking. For each disease, probes were ranked in the development set by TreeSHAP importance relative to noise features over 40 iterations. Probes were retained if they exceeded a null-calibrated stability threshold and a median signal-to-noise cut-off. The bars are schematic. (D) Survival models. Ten-year models were fitted within the development set using five-fold cross-validation. The CoxPHEnsemble comprises 100 penalised Cox members per fold (500 in total), and its risks are averaged to give the MRS. Elastic-net Cox and extra survival trees were also fitted. Penalties were tuned on the validation folds, and the holdout was not used for fitting and tuning. (E), Holdout evaluation at 10 years: discrimination (inverse-probability-of-censoring-weighted C-index and cumulative/dynamic AUC, with 95% bootstrap confidence intervals from 10,000 resamples), hazard ratio per standard deviation of the score (age- and sex-adjusted and unadjusted), closed-form SHAP feature attribution, and pathway and trait enrichment of the panel CpGs. The curves and icons are schematic.

**Table 1.** Cohort characteristics.

| Cohort | Partition | n | Age, mean (SD) | Female % | Control follow-up, median years |
|---|---|---|---|---|---|
| GS | All | 10,156 | 49.3 (15.4) | 56.1 | 15.1 - 15.2 |
|  | Training | 7,109 | 49.3 (15.3) | 56.1 | - |
|  | Validation | 1,522 | 49.1 (15.3) | 56.1 | - |
|  | Holdout | 1,525 | 49.3 (15.6) | 56.1 | - |
| MGB | All | 2,629 | 57.2 (16.0) | 64.1 | 6.6 - 6.8 |
|  | Training | 1,836 | 57.1 (15.9) | 64.0 |  |
|  | Validation | 398 | 57.3 (16.7) | 64.1 |  |
|  | Holdout | 395 | 57.5 (15.8) | 64.3 |  |
| All | All | 12,785 | 50.9 (15.8) | 57.7 | - |

*Age is at sample collection. GS, Generation Scotland; MGB, Mass General Brigham; SD, standard deviation. Samples were assigned to training, validation and holdout partitions (70/15/15) at the sample level, before any feature selection; training and validation were pooled as the development set (n = 10,865). Control follow-up is the median years from sample collection to the end of the follow-up among controls, given as the range across the 21 diseases. Each participant contributes one sample.*

**Table 2.** Cases and controls per disease.

| Disease | Development cases / controls | Holdout cases / controls (any follow-up) | Holdout events by 10y |
|---|---|---|---|
| CKD | 607 / 9,660 | 111 / 1,705 | 92 |
| PVD | 388 / 9,938 | 59 / 1,769 | 59 |
| COPD | 628 / 9,548 | 109 / 1,700 | 86 |
| CHF | 592 / 9,662 | 120 / 1,688 | 95 |
| T2DM | 854 / 9,486 | 133 / 1,701 | 109 |
| Prostate cancer* | 265 / 10,481 | 47 / 738 | 29 |
| ILD | 227 / 10,441 | 44 / 1,832 | 40 |
| Dementia (non-specific) | 316 / 10,528 | 55 / 1,862 | 40 |
| NAFLD | 340 / 10,223 | 56 / 1,813 | 49 |
| Breast cancer* | 332 / 10,211 | 81 / 986 | 59 |
| CAD | 1,160 / 8,440 | 204 / 1,474 | 168 |
| Lung cancer | 236 / 10,547 | 47 / 1,864 | 36 |
| Stroke | 582 / 9,959 | 125 / 1,737 | 104 |
| MI | 544 / 9,727 | 98 / 1,711 | 81 |
| RA | 201 / 10,347 | 36 / 1,820 | 28 |
| Depression | 743 / 8,438 | 141 / 1,521 | 133 |
| Allergic rhinitis | 555 / 8,973 | 100 / 1,582 | 93 |
| Colorectal cancer | 233 / 10,520 | 48 / 1,854 | 33 |
| Asthma | 578 / 8,261 | 114 / 1,460 | 99 |
| Psoriasis | 205 / 10,309 | 39 / 1,808 | 38 |
| Alcohol-related liver disease | 526 / 9,986 | 93 / 1,760 | 83 |

*Development = training + validation pooled. Prevalent cases at sample collection were excluded for each disease. Cases developed disease at any time during follow-up; events are the cases diagnosed within 10 years of collection. CHF, congestive heart failure; CKD, chronic kidney disease; COPD, chronic obstructive pulmonary disease; CV, cross-validation; ILD, interstitial lung disease; NAFLD, non-alcoholic fatty liver disease; PAD, peripheral artery disease; PVD, peripheral vascular disease; T2DM, type 2 diabetes. MI, myocardial infarction; RA, rheumatoid arthritis; CAD, coronary artery disease. \*Holdout controls for prostate and breast are within-sex.*

### Eight of 21 scores reach a 10-year C-index of at least 0.80

Holdout C-index ranged from 0.888 (CKD) to 0.619 (alcohol-related liver disease) (Figure 2A, Table 3). Eight scores reached at least 0.80: CKD (0.888), PVD (0.870), COPD (0.864), CHF (0.843) and T2DM (0.809), each with at least 50 holdout events (tier 1), and ILD, dementia and NAFLD/MASLD (0.842-0.867), each with fewer events and wider intervals (tier 1b). Seven scores fell between 0.70 and 0.80 (tier 2: prostate cancer, coronary artery disease, lung cancer, stroke, myocardial infarction, rheumatoid arthritis, depression) and six below 0.70 (tier 3: breast cancer, allergic rhinitis, colorectal cancer, asthma, psoriasis, alcohol-related liver disease; 0.619-0.695). For alcohol-related liver disease the 10-year AUC was 0.565 (95% CI 0.494-0.636), so it did not discriminate at that horizon. For prostate cancer and breast cancer we report the performance metrics calculated on a within-sex holdout.

**Figure 2.**
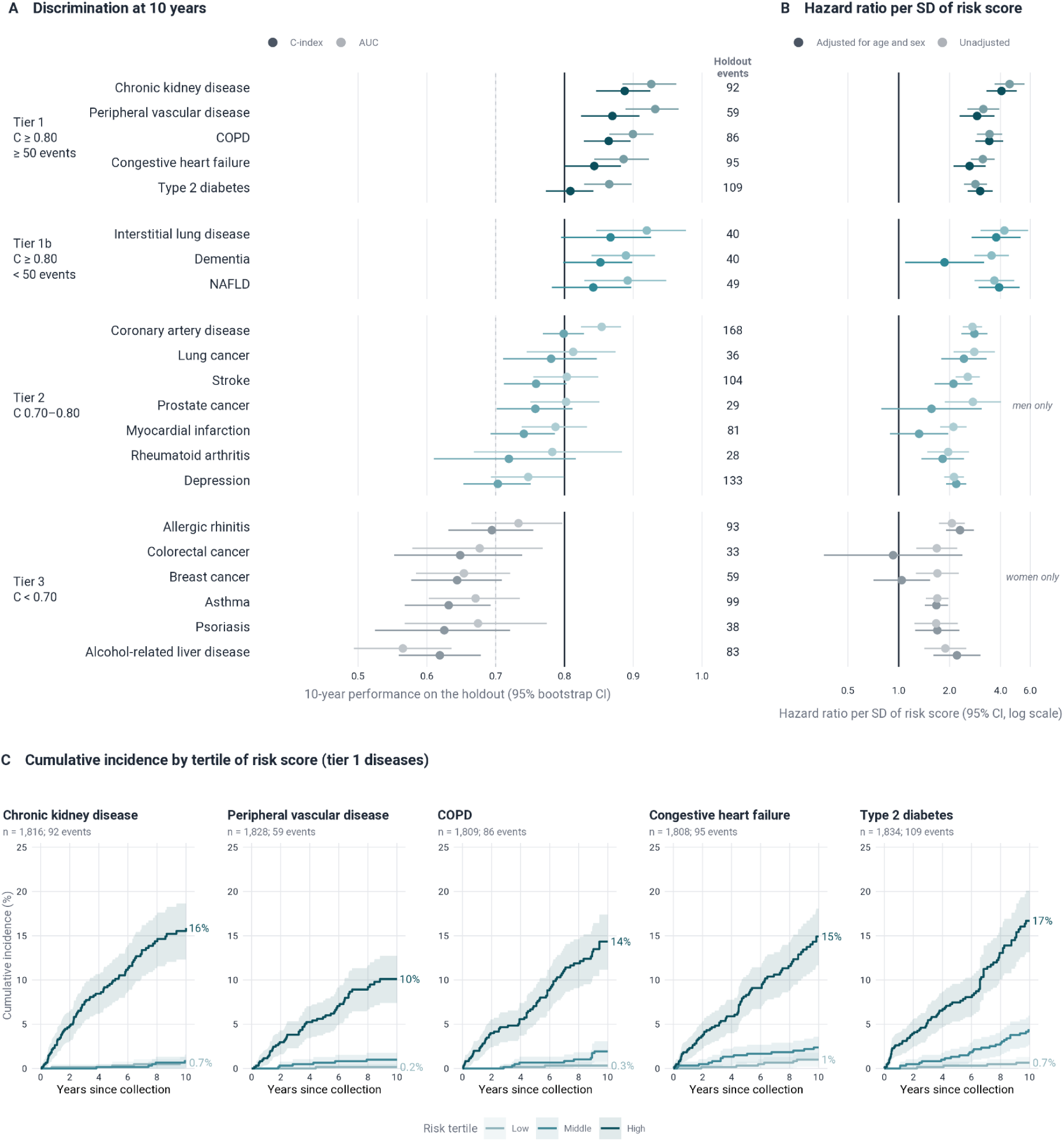
Discrimination and hazard ratios of the 21 methylation risk scores and cumulative incidence of five diseases by tertile of model risk score in the holdout set. (A) Ten-year C-index (inverse-probability-of-censoring-weighted, truncated at 10 years) and cumulative/dynamic area under the curve (AUC) for each disease, with 95% bootstrap confidence intervals (10,000 resamples of the holdout). Diseases are grouped into tiers by the unrounded C-index and the number of holdout events: tier 1, C-index at least 0.80 with at least 50 events; tier 1b, at least 0.80 with fewer than 50 events; tier 2, 0.70 to below 0.80; tier 3, below 0.70. Solid vertical line, C-index 0.80; dashed line, 0.70. The number to the right of each row is the number of holdout events within 10 years. (B) Hazard ratio per standard deviation of the 10-year risk score, from a Cox proportional hazards model fitted in the holdout over full follow-up, adjusted for age and sex (dark) or unadjusted (light), with 95% confidence intervals; log scale, reference line at 1. Rows are aligned with panel A. Prostate and breast cancer occur in one sex, so their C-index, AUC and hazard ratios are for the at-risk sex only (men, 785, 47 events; women, 1,067, 81 events; scores not retrained within sex) and the adjusted value is adjusted for age alone. (C) Kaplan-Meier cumulative incidence over 10 years from sample collection, by tertile of the risk score in the holdout, for the five tier 1 diseases (n = 1,808–1,834 per disease). Tertile cutpoints were computed within the holdout. Follow-up was censored at 10 years or at last follow-up if earlier. Shaded bands are pointwise 95% confidence intervals; values at the right of each curve are the 10-year cumulative incidence in the low and high tertiles. Scores are not calibrated, tertiles are relative to this cohort, and curves are not adjusted for age or sex.

**Table 3.** Selected models, performance and hazard ratios for each disease.

| **Tier** | **Disease** | **Selected model** | **C-index (95% CI)** | **AUC (95% CI)** | **HR per SD, adjusted (95% CI)** | **HR per SD, unadjusted (95% CI)** |
|---|---|---|---|---|---|---|
| **1** | CKD | ExtraSurvivalTrees | 0.888 (0.846-0.925) | 0.926 (0.884-0.963) | 4.06 (3.30-4.99) | 4.53 (3.69-5.55) |
|  | PVD | CoxPHEnsemble | 0.870 (0.824-0.909) | 0.932 (0.889-0.966) | 2.91 (2.29-3.68) | 3.16 (2.55-3.93) |
|  | COPD | CoxPHEnsemble | 0.864 (0.828-0.896) | 0.900 (0.865-0.930) | 3.43 (2.84-4.15) | 3.45 (2.90-4.09) |
|  | CHF | CoxPHEnsemble | 0.843 (0.800-0.882) | 0.886 (0.843-0.923) | 2.62 (2.11-3.27) | 3.14 (2.67-3.70) |
|  | T2DM | CoxPHEnsemble | 0.809 (0.773-0.842) | 0.865 (0.829-0.898) | 3.03 (2.56-3.60) | 2.84 (2.43-3.33) |
| **1b** | ILD | CoxPH | 0.867 (0.795-0.926) | 0.920 (0.846-0.977) | 3.77 (2.70-5.27) | 4.22 (3.05-5.84) |
|  | Dementia (non-specific) | CoxPH | 0.852 (0.798-0.899) | 0.889 (0.839-0.932) | 1.87 (1.09-3.20) | 3.54 (2.80-4.48) |
|  | NAFLD | ExtraSurvivalTrees | 0.842 (0.782-0.897) | 0.892 (0.829-0.948) | 3.93 (2.97-5.19) | 3.68 (2.81-4.82) |
| **2** | CAD | CoxPHEnsemble | 0.799 (0.769-0.829) | 0.854 (0.824-0.882) | 2.80 (2.35-3.35) | 2.73 (2.39-3.11) |
|  | Lung cancer | ExtraSurvivalTrees | 0.781 (0.711-0.847) | 0.813 (0.745-0.874) | 2.43 (1.78-3.31) | 2.80 (2.11-3.71) |
|  | Stroke | CoxPHEnsemble | 0.759 (0.712-0.803) | 0.803 (0.755-0.849) | 2.11 (1.63-2.73) | 2.56 (2.17-3.02) |
|  | Prostate cancer* | ExtraSurvivalTrees | 0.758 (0.701-0.812) | 0.802 (0.750-0.851) | 1.56 (0.79-3.10) | 2.75 (1.87-4.03) |
|  | MI | CoxPHEnsemble | 0.741 (0.693-0.786) | 0.787 (0.738-0.833) | 1.32 (0.89-1.96) | 2.11 (1.76-2.52) |
|  | RA | CoxPH | 0.719 (0.610-0.817) | 0.782 (0.668-0.884) | 1.82 (1.36-2.43) | 1.96 (1.48-2.60) |
|  | Depression | CoxPHEnsemble | 0.703 (0.653-0.751) | 0.747 (0.693-0.799) | 2.19 (1.91-2.51) | 2.12 (1.86-2.43) |
| **3** | Allergic rhinitis | ExtraSurvivalTrees | 0.695 (0.631-0.755) | 0.733 (0.665-0.797) | 2.30 (1.90-2.79) | 2.06 (1.73-2.46) |
|  | Colorectal cancer | CoxPH | 0.648 (0.552-0.739) | 0.677 (0.579-0.768) | 0.93 (0.36-2.38) | 1.68 (1.27-2.22) |
|  | Breast cancer* | ExtraSurvivalTrees | 0.644 (0.577-0.709) | 0.654 (0.584-0.721) | 1.04 (0.71-1.53) | 1.69 (1.26-2.27) |
|  | Asthma | CoxPHEnsemble | 0.632 (0.568-0.693) | 0.671 (0.603-0.735) | 1.67 (1.42-1.96) | 1.69 (1.45-1.97) |
|  | Psoriasis | CoxPH | 0.625 (0.524-0.721) | 0.674 (0.568-0.774) | 1.69 (1.25-2.29) | 1.66 (1.24-2.24) |
|  | Alcohol-related liver disease | ExtraSurvivalTrees | 0.619 (0.559-0.678) | 0.565 (0.494-0.636) | 2.21 (1.60-3.04) | 1.89 (1.42-2.51) |

*AUC, area under the receiver operating characteristic curve; CHF, congestive heart failure; CKD, chronic kidney disease; COPD, chronic obstructive pulmonary disease; CV, cross-validation; ILD, interstitial lung disease; NAFLD, non-alcoholic fatty liver disease; PAD, peripheral artery disease; PVD, peripheral vascular disease; T2DM, type 2 diabetes. MI, myocardial infarction; RA, rheumatoid arthritis; CAD, coronary artery disease. \* Performance metrics are evaluated on a within-sex holdout, with HRs adjusted for age alone.*

For each disease, we calculated hazard ratios per SD with and without adjusting by age and sex. Scores remained associated with disease following adjustment for age and sex for 17 of the 21 diseases (Figure 2B). Adjusted hazard ratios (HR) per SD were 2.62-4.06 in tier 1 (CKD 4.06, 95% CI 3.30-4.99). The greatest attenuation of the association as a result of age- and sex-adjustment was observed for dementia (3.54 to 1.87), MI (2.11 to 1.32, 95% CI 0.89-1.96) and colorectal cancer (1.68 to 0.93, 0.36-2.38), with MI and colorectal cancer becoming statistically non-significant.

Prostate cancer, and the majority of breast cancer cases, occur in one sex, so mixed-sex holdout scores carry a signal that discriminates events largely on sex-associated CpGs (reported at p < 1×10⁻⁷ in blood EWAS^31^: 146 of 380 CpGs, 38%, of the prostate panel and 196 of 478 CpGs, 41%, of the breast panel, against 0.4% of background probes). We therefore re-evaluated both scores within the at-risk sex, without retraining, and compared them with age alone (prostate, 785 men, 47 events over full follow-up; breast, 1,067 women, 81 events). For prostate cancer, the 10-year C-index was 0.758 (AUC 0.802), and the age-adjusted HR per SD was 1.56 (95% CI 0.79-3.10) (Figure 2B). For breast cancer, the C-index was 0.644 (AUC 0.654), with an age-adjusted hazard ratio of 1.04 (95% CI 0.71-1.53). Within sex, the prostate score adds little to age and the breast score does not add to it.

Ranking holdout participants by risk score separated 10-year cumulative incidence clearly in the five tier 1 diseases (Figure 2C). Incidence in the highest tertile was 15.9% for CKD, 10.1% for PVD, 14.3% for COPD, 14.9% for CHF and 16.7% for T2DM, against 0.7%, 0.2%, 0.3%, 1.0% and 0.7% in the lowest. The highest tertile contained 73-90% of all 10-year events (CKD 83 of 92, PVD 52 of 59, COPD 73 of 86, CHF 75 of 95, T2DM 80 of 109). The lowest tertile held only 1-6 events per disease, so low-tertile estimates and the ratios between tertiles are imprecise. Scores are not calibrated, so absolute risks are relative to this cohort. Curves for the other 16 diseases are in Supplementary Figure 1.

### Holdout performance alongside published methylation and polygenic scores

We set our holdout estimates beside published values for the 21 diseases (Figure 3 and Table 4 for the eight tier 1 and 1b diseases; all 125 studies in Supplementary Tables 2 and 3 and Supplementary Figure 2). These are values identified in a targeted search and reported from other cohorts, horizons and designs, not a head-to-head comparison. Of the 125 published studies (62 methylation score and 63 polygenic score studies), 53 concerned the eight tier 1 and 1b diseases, and 49 gave at least one closest comparison, that is, the same disease, an AUC or C-index and, for methylation scores, an incident design (15 methylation, 34 polygenic; 25 for the tier 1 and 1b diseases). The rest were context comparisons or not comparable (for example prevalent or cross-sectional methylation scores, general-purpose epigenetic clocks, or a different outcome).

**Figure 3.**
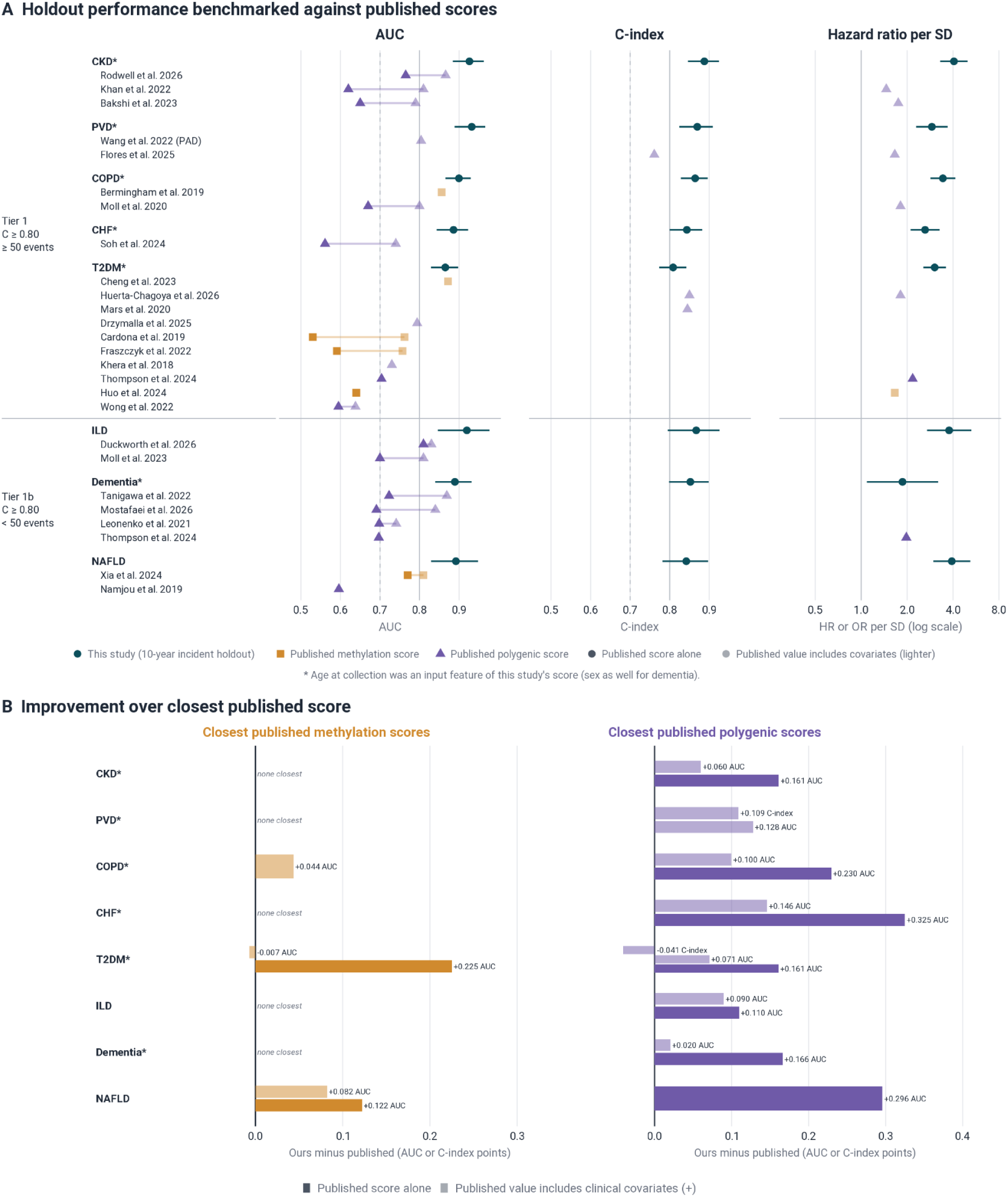
Benchmark of the methylation risk scores against published methylation and polygenic risk scores for tier 1 and tier 1b. (A) Holdout AUC, C-index and hazard ratio per standard deviation (SD) of our 10-year methylation risk scores (dark circles; 95% bootstrap confidence intervals for AUC and C-index; 95% confidence intervals for hazard ratios from Cox models adjusted for age and sex, log scale) beside the closest published scores for the eight tier 1 and 1b diseases. Each block is one disease: the first row is this study and each further row is one published study. Where a study reports several values (different cohorts, scores or subgroups), only the highest is shown, separately for score alone and with covariates; all values are in Supplementary Tables 2 and 3. Only closest comparisons are shown: the same disease, an AUC or C-index and, for methylation scores (orange squares), an incident design. Polygenic scores (purple triangles) from case-control or prevalent-case designs are also included, because genotype is fixed at birth. Published values are point estimates as reported. Darker symbols are scores alone; lighter symbols are models that also include covariates (for example age, sex, principal components or clinical factors). Joined points are several values from one study. Published hazard or odds ratios are as reported. Solid line, 0.80; dashed line, 0.70. Published follow-up horizons differ from our 10 years. Polygenic scores for interstitial lung disease (ILD) are for idiopathic pulmonary fibrosis, a subtype. (B) Our holdout value minus the highest closest published value (AUC points or, where only a C-index was published, C-index points), for methylation scores (orange) and polygenic scores (purple), separately for published scores alone (darker) and values including covariates (lighter). “None closest” means no published score met the closest definition. The negative bars (type 2 diabetes) are published values higher than ours. Published values estimated in the data used to select the score (Mars et al.^5^, evaluated in or overlapping the cohort used to choose the tuning parameter; Cardona et al.^32^, Xia et al.^33^ and Huo et al.^34^ apparent estimates) are optimistic, so these differences are conservative (marked † in Table 4). * Panel includes age at collection as a feature.

**Table 4.** Holdout performance of our MRS (10-year incident disease) alongside the closest published methylation scores and polygenic scores.

| Tier | Disease | Our C-index / AUC | Published MRS, same endpoint |  | Published PRS, same endpoint |  |
|---|---|---|---|---|---|---|
|  |  |  | Score alone | +covariates | Score alone | +covariates |
| **1** | CKD | 0.888 / 0.926 | *None for this endpoint*<br>Nearest: Marchiori et al.<sup>40</sup> (T2DM population only) AUC 0.82 | Nearest: Marchiori et al.<sup>40</sup> AUC 0.87 (+ clinical) | **AUC 0.765** (prevalent; Rodwell et al.<sup>82</sup>)<br>**AUC 0.65** (prevalent; Bakshi et al.<sup>83</sup>)<br>**AUC 0.62** (prevalent; Khan et al.<sup>58</sup>) | **AUC 0.866** (+ clinical; prevalent; Rodwell et al.<sup>82</sup>)<br>**AUC 0.81** (+ clinical; prevalent; Khan et al.<sup>58</sup>)<br>**AUC 0.79** (+ clinical; prevalent; Bakshi et al.<sup>83</sup>) |
|  | PVD | 0.870 / 0.932 | *None for this endpoint* | Nearest: Barad et al.<sup>25</sup> (preprint) C-index 0.760 (+ clinical) | — | **AUC 0.804** (+ clinical; prevalent; Wang et al.<sup>84</sup>)<br>**C-index 0.761** (+ covariates; Flores et al.59**)** |
|  | COPD | 0.864 / 0.900 | — | **AUC 0.856** (+ clinical; Bermingham et al.<sup>49</sup>) | **AUC 0.67** (prevalent; Moll et al.<sup>60</sup>) | **AUC 0.80** (+ clinical; prevalent; Moll et al.<sup>60</sup>) |
|  | CHF | 0.843 / 0.886 | *None for this endpoint*<br>Nearest: Zhao et al.<sup>42</sup> (HFpEF only) AUC 0.65 | Nearest: Zhao et al.<sup>42</sup> (HFpEF only) AUC 0.90 (+ clinical) | **AUC 0.561** (Soh et al.<sup>51</sup>) | **AUC 0.74** (+ covariates; Soh et al.<sup>51</sup>) |
|  | T2DM | 0.809 / 0.865 | **AUC 0.64** (Huo et al.<sup>34</sup> †)<br>**AUC 0.591** (Fraszczyk et al.<sup>50</sup>)<br>**AUC 0.53** (Cardona et al.<sup>32</sup> †) | **AUC 0.872** (+ clinical, protein epigenetic score; Cheng et al.<sup>22</sup>)<br>**AUC 0.762** (+ clinical; Cardona et al.<sup>32</sup> †)<br>**AUC 0.757** (+ clinical; Fraszczyk et al.<sup>50</sup>) | **AUC 0.704** (prevalent; Thompson et al.<sup>6</sup>)<br>**AUC 0.595** (Wong et al.<sup>85</sup>) | **C-index 0.85** (+ clinical; Huerta-Chagoya et al.<sup>35</sup>)<br>**C-index 0.845** (+ clinical; Mars et al.<sup>5</sup> †)<br>**AUC 0.794** (+ clinical; prevalent; Drzymalla et al.<sup>53</sup>)<br>**AUC 0.73** (+ covariates; prevalent; Khera et al.<sup>3</sup>)<br>**AUC 0.638** (+ covariates; Wong et al.<sup>85</sup>) |
| **1b** | ILD | 0.867 / 0.920 | *None for this endpoint* | — | **AUC 0.81** (prevalent; Duckworth et al.<sup>52</sup>)<br>**AUC 0.70** (prevalent; Moll et al.<sup>54</sup>) | **AUC 0.83** (+ covariates; prevalent; Duckworth et al.<sup>52</sup>)<br>**AUC 0.81** (+ clinical; prevalent; Moll et al.<sup>54</sup>) |
|  | Dementia (non-specific) | 0.852 / 0.889 | *None for this endpoint* | Nearest: Hackenhaar et al.<sup>48</sup> (case-control) AUC 0.78 (+ clinical) | **AUC 0.723** (prevalent; Tanigawa et al.<sup>39</sup>)<br>**AUC 0.698** (prevalent; Leonenko et al.<sup>55</sup>)<br>**AUC 0.697** (prevalent; Thompson et al.<sup>6</sup>)<br>**AUC 0.691** (Mostafaei et al.<sup>56</sup>) | **AUC 0.869** (+ covariates; prevalent; Tanigawa et al.<sup>39</sup>)<br>**AUC 0.83** (+ covariates; Mostafaei et al.<sup>56</sup>)<br>**AUC 0.741** (+ clinical; prevalent; Leonenko et al.<sup>55</sup>) |
|  | NAFLD | 0.842 / 0.892 | **AUC 0.77** (Xia et al.<sup>33</sup> †) | **AUC 0.81** (+ clinical; Xia et al.<sup>33</sup> †) | **AUC 0.596** (prevalent; Namjou et al.<sup>86</sup>) | — |

*Published values are shown as reported, C-index first and AUC second. Where a study reports several values (different cohorts, scores or subgroups), only the highest is shown, separately for score alone and with covariates; all reported values are in Supplementary Tables 2 and 3. “Score alone” means no covariates; “+ covariates” means demographic, technical or genetic covariates only (age, sex, principal components, array, batch, cell types); “+ clinical” means any clinical or biological risk factor or risk score (for example BMI, lipids, blood pressure, glucose, diabetes, smoking, family history, or APOE genotype), as in Supplementary Tables 2 and 3. “Prevalent” marks polygenic score values estimated in case-control, case-status or prevalent-case designs, which are accepted because genotype is fixed at birth; all methylation values are from incident designs. † Estimate possibly derived in the data used to select the score, CpGs, weights or tuning parameter, so the published value may be optimistic and the comparison conservative (see Supplementary Tables 2 and 3). “None for this endpoint” means no published score met the closest criteria for the disease (same disease, an AUC or C-index and, for methylation scores, an incident design); the nearest context comparison is shown in italics. —, no published value. The published ILD polygenic scores are for idiopathic pulmonary fibrosis (IPF), a subtype of ILD. Tanigawa et al.<sup>39</sup>, Leonenko et al.<sup>55</sup> and Thompson et al.<sup>6</sup> report Alzheimer’s disease for dementia. Mostafaei et al.<sup>56</sup> reports Alzheimer’s disease and related dementias.*

Across the eight tier 1 and 1b diseases, our MRS scored higher than 64 of the 68 closest published AUC or C-index values (25 studies). Across all 21 diseases, using the within-sex values for prostate and breast cancer, our MRS scored higher than 116 of the 140 closest published values (49 studies) and was the highest in 11 of the 18 diseases that have a closest comparator. The 24 exceptions fall in seven diseases: T2DM (four values from three studies that added clinical risk factors to the score, including polygenic C-indices of 0.85 and 0.845 against our 0.809)^5,22,35^, lung cancer (a methylation score alone, AUC 0.85, and a methylation score with clinical factors, 0.817, against our 0.813)^23,36^, colorectal cancer (two methylation scores, AUC 0.82, with CpGs possibly screened before the train-test split, and 0.702, against our 0.677)^37,38^, psoriasis (four polygenic values, 0.685 to 0.702 against our 0.674)^6,39^ and CAD (polygenic C-indices of 0.832 and 0.820 against our 0.799, evaluated in the cohort used to choose the tuning parameter)^5^, prostate cancer (polygenic C-indices of 0.857 and 0.866,^5^ and AUCs of 0.833 and 0.836, against our within-sex C-index of 0.758 and AUC of 0.802) and breast cancer (four AUCs of 0.666 to 0.71 and two polygenic C-indices of 0.737 and 0.750, against our within-sex AUC of 0.654 and C-index of 0.644).^5,6,24^ Ten of the 49 studies may have been estimated in the data used to select the score, CpGs, weights or tuning parameter, so their values may be optimistic; these are marked as exceptions in Supplementary Tables 2 and 3 and with † in Table 4.

Among the eight diseases only one published methylation score exceeded ours, and only marginally. Cheng et al.^22^ reached an AUC of 0.872 for T2DM against our 0.865. Their model combines clinical risk factors (which alone reach 0.839) with two methylation scores, unlike our methylation score, which uses methylation and age only. More importantly, to our knowledge, we report 10-year incident MRS for diseases with no close comparator: this includes five of the eight (CKD, PVD, CHF, ILD, dementia), and 14 of all 21, where no published methylation score with the same endpoint and an incident design exists (Supplementary Table 2). The nearest are a methylation score for CKD in people with T2DM only,^40^ composite cardiovascular scores,^25,41^ heart failure with preserved ejection fraction,^42^ and case-control or case-only classifiers for prostate cancer,^43,44^ ILD^45^ and dementia.^46–48^ Where a published comparator exists, our MRS exceeds it in most cases (Figure 3B). For COPD,^49^ our AUC of 0.900 was 0.04 above a published score that includes clinical factors (0.856); for T2DM, 0.865 was 0.23–0.27 above published scores alone (0.591–0.64);^34,50^ and for NAFLD,^33^ 0.892 was 0.12 above a published score alone (0.77) and 0.08 above the score with clinical factors (0.81), although those estimates come from the same participants used to select the CpGs.

Our MRS also exceeded reported values for PRS alone for every disease that has one, by 0.11 to 0.33 AUC (ILD +0.11, CKD +0.16, T2DM +0.16, dementia +0.17, COPD +0.23, NAFLD +0.30, CHF +0.33), against published AUCs of 0.56 to 0.81.^51,52^ Adding age, sex, principal components or clinical factors lifted polygenic models to AUCs of 0.64 to 0.87. Comparing AUC and C-index, ours remained higher for six of the seven diseases with such a value (AUC against AUC, C-index against C-index), by +0.02 AUC (dementia), +0.06 AUC (CKD), +0.09 AUC (ILD), +0.10 AUC (COPD), +0.11 C-index and +0.13 AUC (PVD) and +0.15 AUC (CHF). The exception is T2DM, where a clinical risk score combined with a polygenic score reached a C-index of 0.85 against our 0.809, although our AUC (0.865) exceeded the best published polygenic AUC (0.794, +0.07).^53^ Most polygenic values come from prevalent case-control or mixed prevalent and incident analyses, largely in UK Biobank,^6^ and describe lifetime rather than 10-year risk, so they are not directly comparable. The ILD scores are for idiopathic pulmonary fibrosis,^52,54^ a subtype of ILD. The dementia scores are for Alzheimer’s disease or Alzheimer’s disease and related dementias.^6,39,55,56^

Taken together, ours was higher than the closest published polygenic or methylation score that included covariates for seven of the eight diseases; the exception was T2DM (AUC 0.872 against our 0.865 for a methylation score with clinical risk factors, and C-index 0.85 against our 0.809 for a polygenic score with clinical risk factors).

### Panel CpGs map to inflammatory, sterol-metabolism and synaptic gene sets

We tested gene-set over-representation among panel CpGs for all 21 diseases (gene sets of at most 1,000 genes, FDR < 0.05 within disease and collection). Eight diseases had at least one significant set and 13 had none, including CKD, PVD, ILD and NAFLD. The clearest results were for four diseases: COPD (7 sets), CHF (4), T2DM (5) and dementia (4) (Figure 4A); prostate cancer (10 sets, mainly KEGG neurodegeneration pathways), breast cancer (1), asthma (1) and CAD (1) each had sets at FDR < 0.05 that are not shown in Figure 4. Sixteen of these 20 sets remained significant with all EPICv1 probes as the background. The sets fell into distinct themes. For COPD they were cytokine signalling, led by TNF-α signalling via NF-κB (FDR 4.4 × 10⁻⁴; 13 panel genes), with IL6-JAK-STAT3 (0.0069) and IL2-STAT5 (0.022). For CHF they were innate immune sensing: NLR signalling (FDR 0.0042; 8 genes), SARS-CoV-1 modulation of innate immune responses (0.016), inflammasomes (0.019) and interferon-α response (0.021). For T2DM they were TNF-α signalling (FDR 3.0 × 10⁻⁴; 13 genes) and sterol metabolism, including cholesterol metabolism (Bloch and Kandutsch-Russell pathways; 0.0026; 7 genes) and zymostenol biosynthesis (0.0049; 4 genes). For dementia they were four overlapping synaptic signalling terms (FDR 0.028-0.031; 26 genes each), which count as one finding. No gene set reached FDR < 0.05 for PVD or CKD. Restricting the panels to their top 25% or 50% of CpGs by SHAP weight kept TNF-α signalling in COPD (FDR 0.007 at the top 25%) and the cholesterol and zymostenol sets in T2DM (0.0006 and 0.0057), but not the innate-immune sets for CHF or the synaptic terms for dementia (all FDR > 0.05), so those two findings depend on which CpGs are in the panel more than on the most heavily weighted ones.

**Figure 4.**
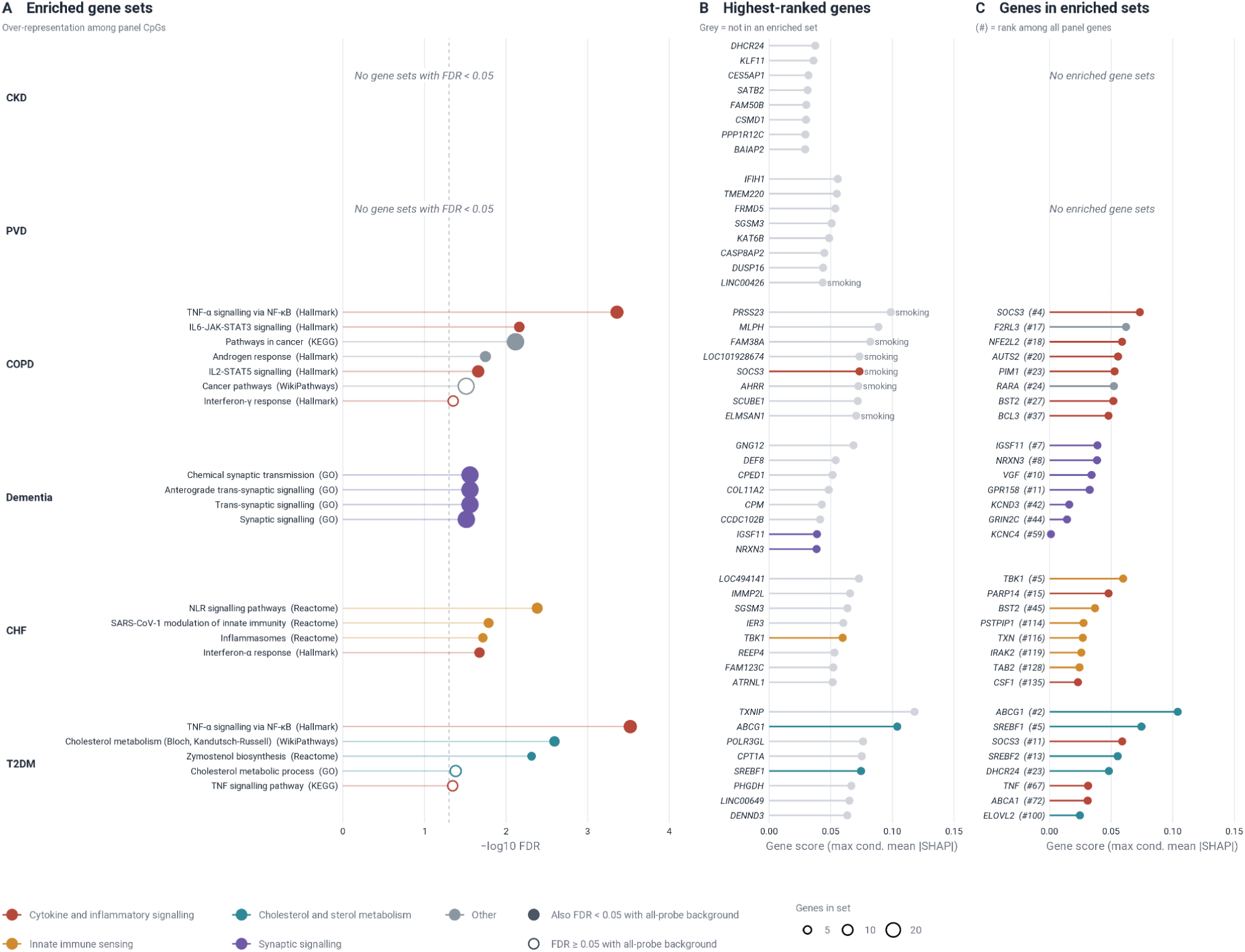
Gene sets and genes behind the scores with significant enrichment. (A) Gene sets over-represented among panel CpGs for the six tier 1 and 1b diseases (chronic kidney disease, peripheral vascular disease, COPD, dementia, congestive heart failure, type 2 diabetes; ordered by holdout C-index). Sets with FDR < 0.05 (Benjamini-Hochberg within disease and collection; background, the 439,530 probes of the variance-filtered pool; sets of more than 1,000 genes excluded) are shown; none reached FDR < 0.05 for chronic kidney disease or peripheral vascular disease. Colour is the family of the gene set and point size the number of panel genes in the set. Filled points were also FDR < 0.05 with all EPICv1 probes as background. The dashed line marks FDR = 0.05. The four synaptic dementia terms share most genes and are one finding. (B) The eight highest-ranked panel genes for each disease (Illumina hg19 RefGene symbols; CpGs without a hg19 gene are not ranked). Gene score is the largest mean absolute SHAP value, conditional on inclusion, among its CpGs sampled by at least 5 of the 500 ensemble members. Colour is the family of the gene’s most significant enriched set; grey, none. “Smoking” marks genes with a CpG in published whole-blood smoking EWAS (EWAS Atlas, P < 1 × 10⁻⁷), shown for COPD and peripheral vascular disease only. (C) The eight highest-ranked panel genes within each disease’s significant sets, scored as in B; the number in brackets is the gene’s rank among all panel genes. Only sets matched to MSigDB lists (Hallmark, Reactome, WikiPathways, GO synaptic signalling) are shown. SHAP values describe model use of a feature, not the direction or causality of an association.

The genes the models relied on most (Figure 4B) lined up with several of these themes (Figure 4C; ranks are among all panel genes, as in the figure). In T2DM, the two highest-ranked genes were TXNIP (cg19693031; rank 1) and ABCG1 (cg16740586; rank 2), both loci reported in blood EWAS of T2DM.^32,50,57^ The ABCG1 gene sits in the sterol sets together with SREBF1 (rank 5), SREBF2 (13), DHCR24 (23) and ABCA1 (72), and SOCS3 (11) and TNF (67) in the TNF-α set. In COPD, the highest-ranked gene was PRSS23 (rank 1) and the highest-ranked CpG overall, cg06644428, has no gene in the hg19 annotation (GENCODE v36 maps it to the pseudogene ECEL1P1); it is a smoking-associated locus, as is the AHRR CpG (cg05575921; gene rank 6).^9^ SOCS3 (rank 4) was the highest-ranked gene inside an enriched set, followed by F2RL3 (17) and NFE2L2 (18). In CHF the two highest-ranked protein-coding genes (IMMP2L, rank 2; SGSM3, rank 3) were not in an enriched set, whereas TBK1 (rank 5) was in the innate-immune sets, followed by PARP14 (15) and BST2 (45). In dementia, which the selected model fits with 78 of the 258 panel CpGs, the highest-ranked genes were GNG12, DEF8 and CPED1, none in an enriched set; the synaptic genes ranked IGSF11 (7), NRXN3 (8), VGF (10) and GPR158 (11) of 61. For PVD and CKD, which had no enriched sets, the top genes were IFIH1, TMEM220, FRMD5, SGSM3 and KAT6B (PVD) and DHCR24, KLF11, CES5AP1, SATB2 and FAM50B (CKD); none belongs to an enriched gene set.

For two of our MRS models, T2DM and COPD, the CpGs in the model panels coincided with published findings, but not for the other diseases tested. Notably for T2DM, 21 of the 76 CpGs reported by an incident-disease epigenome-wide meta-analysis of five European cohorts (1,250 incident cases and 1,950 controls) were in our 458-CpG model panel, and 21 of its 65 genes were also panel genes.^50^ The overlap included our highest-ranked CpG, TXNIP cg19693031 (rank 1 of 458), two of the three ABCG1 CpGs (ranks 3 and 4), an SREBF1 CpG (rank 8), PHGDH (rank 12), DHCR24 (rank 33) and TLR9 (rank 95). Eight of the 21 were among our 15 highest-ranked CpGs. Importantly, those cohorts (Doetinchem, ESTHER, KORA1, KORA2 and EPIC-Norfolk) are independent of ours, and 64 of the 76 CpGs replicated at nominal significance (p < 0.05) in an Indian Asian cohort (LOLIPOP). For COPD,^49^ 7 of the 28 CpGs reported in a cross-sectional EWAS study were in the model panel, including three SOCS3 CpGs (ranks 6, 9 and 394), AHRR (rank 7), ELMSAN1 (11), F2RL3 (21) and SLAMF7 (155), however this study is not independent of our training data.

Panels also contained many CpGs associated with age in published blood EWAS (EWAS Atlas, P < 1e-7). Seventy-nine percent of the dementia panel (203 of 258 CpGs), 53% for CHF (241 of 458), 33% for T2DM (150 of 458), 27% for COPD (154 of 564), 21% for PVD (45 of 216) and 11% for CKD (38 of 361) were associated with age, compared with 3.0% of the background probes (all FDR < 1e-8). Smoking-associated CpGs made up two thirds of the COPD panel (373 of 564, 66%, against 2.0% of background probes; FDR < 1e-300) and a fifth of the PVD (49 of 216, 23%). These sets are the union of CpGs reported in any blood study at P < 1e-7, so overlap means a published association, not independent confirmation. Panels contained almost none of the CpGs used to deconvolve 12 blood cell types;^11^ none for CKD, PVD, COPD, CHF, prostate cancer, ILD, dementia or NAFLD, and one for T2DM (FDR = 1). Asthma, a tier 3 disease, was the exception (13 of 450 CpGs, 37.5-fold, FDR 1.8 × 10⁻¹⁵), and all 13 were eosinophil marker CpGs. The asthma panel therefore contains an eosinophil signature that no other panel shows. Apart from the asthma, the scores are not built from cell-type marker CpGs, but this does not rule out confounding through other CpGs, and cell composition was not modelled. SHAP values describe how much a model relies on a CpG, not the direction or causality of its association with disease.

## DISCUSSION

We trained ten-year methylation risk scores for 21 common diseases on a single development set drawn from two cohorts and evaluated all of them in a single holdout set aside before any analysis. Eight scores reached a C-index of at least 0.80, with the strongest signal in cardiometabolic and respiratory disease: CKD, PVD, COPD, CHF and T2DM each had at least 50 holdout events, age- and sex-adjusted hazard ratios of 2.6 to 4.1 per standard deviation, and a highest risk tertile that contained 73 to 90% of ten-year events. The same evaluation also showed where blood methylation carries little disease-specific signal at this sample size, in allergic and immune-mediated disease, colorectal cancer and alcohol-related liver disease. Because every score was built and tested the same way, these differences more plausibly reflect the disease pathophysiologies and their representations rather than the methods, which earlier single-disease studies could not establish.^19,20,22,49^

Set beside published values, the holdout performance compares well. For every top-tier disease with a published polygenic score alone (seven of the eight), our scores outperformed it, despite using no clinical risk factors. They also outperformed polygenic or methylation scores combined with age, sex or clinical factors for seven of the eight diseases.^6,39,51,52,54–56,58–60^ The exception is T2DM, where polygenic^5,35^ and methylation^22^ models that add clinical risk factors had higher values. Published methylation scores with the same endpoint and an incident design exist for only three of these diseases (COPD, T2DM and NAFLD), and ours were higher except for those T2DM models.^22,33,49^ For the other five top-tier diseases (CKD, PVD, CHF, ILD and dementia; 14 of the 21 diseases overall) we found no published incident methylation score with an AUC or C-index for the same endpoint. The T2DM comparison is the closest, because it uses the same cohort. Our score, without clinical risk factors, was marginally below the published model that combines clinical risk factors with methylation in GS,^22^ and our AUC (0.865) exceeded the best published polygenic AUC (0.794).^53^ These comparisons are of reported values across different cohorts, horizons and case definitions, not head-to-head analyses in the same participants. Most published PRS discrimination comes from prevalent or mixed case-control designs,^60,61^ largely in UK Biobank,^6^ whereas ours is ten-year incident risk in a holdout. The direction, that methylation discriminates better than genotype for exposure- and metabolism-driven diseases, is nonetheless consistent with the one earlier direct comparison, in which methylation scores outperformed polygenic scores for most phenotypes in 831 patients,^26^ and with the view that the two carry complementary information.^62^

The CpGs the models rely on are consistent with known biology and suggest sources of the signal. The T2DM score draws on TXNIP and ABCG1, the most replicated loci in blood methylation studies of the disease, and its panel is enriched for TNF-α signalling and sterol metabolism; the COPD panel is enriched for cytokine signalling and contains CpGs from the smoking axis (AHRR, F2RL3, PRSS23); the heart failure panel is enriched for innate immune sensing. Age and sex adjustment changed the hazard ratios for the cardiometabolic and respiratory scores modestly, reduced them by about half or more for myocardial infarction and dementia, and removed the association for colorectal cancer. Moreover, 79% of the dementia panel consists of CpGs associated with age in published studies. Two conclusions follow. For the strongest scores, the signal largely persists after adjustment for age and sex and is plausibly a readout of inflammatory, metabolic and exposure state. For dementia, and to a lesser extent the other tier 1b diseases, part of the discrimination is age, and the adjusted estimates should be read as the relevant ones. For prostate and breast cancer, which occur in one sex, the mixed-sex values partly reflect sex-associated CpGs, so the within-sex values are the relevant ones. SHAP values describe how a model uses a CpG and not the direction or causality of its association.

The clinical argument for these scores rests on the breadth of information available from a single methylation profile. The 21 scores are computed from a single whole-blood assay; the same assay which supports multiple methylation-based measures that have been developed over the past decade. These include epigenetic clocks of biological age and pace of ageing,^63–65^ scores that estimate circulating protein and metabolite levels,^12,16^ predictors of smoking, alcohol and body mass,^10^ and reference-based deconvolution of twelve immune subtypes.^11^ Platforms that compute many of these measures from one array already exist.^66^ A clinician ordering one test could therefore receive ten-year risk ranks for several diseases, alongside the biological age, immune and biomarker context needed to interpret them, akin to a metabolic panel read as a whole rather than as isolated values. Polygenic risk scores cannot provide this, because they are fixed at conception, report lifetime liability rather than current state, and have shown clinical validity but not clinical utility.^2,4,7^ A methylation profile changes with exposure and intervention,^9,13^ so the same test can, in principle, be repeated to show whether risk has moved. We have not tested that here, and longitudinal evaluation of these scores is the most important next step. Moreover, accessibility matters as much as accuracy for a test meant to be repeated. Methylation arrays run on dried blood spots give profiles highly concordant with venous blood on the 450K array from the same individuals,^67^ the same BeadChip technology as the EPIC array used here, so a card collected at home or in a primary-care clinic could in principle be mailed at ambient temperature and run on the same platform. This could widen the population that can be stratified and followed to people for whom phlebotomy, specialist referral or genotyping is impractical, which is where ten-year risk stratification for chronic disease may have the most to offer.

This study has limitations. The two cohorts differ in setting, age, outcome ascertainment and follow-up. Although the partitions were stratified by cohort, feature selection and the null threshold were pooled across the two cohorts rather than derived in each cohort separately, so we did not test whether cohort-specific null models, which could differ with demographics and outcome ascertainment, would select different CpGs, and transfer of each score from one cohort to the other has not been tested. GS is a family-based cohort and partitions were assigned at the individual level, so relatives may be divided between the development and holdout sets, which could inflate holdout performance. Scores were fitted on class-balanced training folds and are not population-calibrated, so they rank individuals but do not yet give absolute risks; recalibration of the baseline hazard is required before any absolute-risk use. Moreover, the reported model for each disease was the best of three by holdout C-index, so the holdout estimates are somewhat optimistic. Several diseases, including all of tier 1b, had fewer than 50 holdout events, and their confidence intervals are wide. Cell-type composition was not modelled, but when we intersected every panel with blood cell-type marker CpGs, only the asthma panel was enriched for eosinophil markers. This is consistent with asthma biology, as eosinophilic type 2 airway inflammation is a major asthma endotype. Blood EWAS of asthma has also reported eosinophil-linked CpGs.^68^ Both cohorts are predominantly of European ancestry and all samples were measured on one array version, so performance in other ancestries and on newer platforms is unknown. The comparisons with published scores are of reported values rather than head-to-head analyses, as noted above. In particular, we did not benchmark the scores against age, sex and established clinical risk models in the same participants, so their incremental value over routinely available predictors remains to be established. Prevalent disease was excluded on the basis of recorded diagnoses, so undiagnosed disease at sampling may contribute to the prediction of early events. The 2,233 GS participants assigned a fixed censoring time are event-free at ten years by construction. Age at collection was an input to 15 of the 21 scores, including six of the eight top-tier diseases (CKD, PVD, COPD, CHF, T2DM, and dementia), and sex was also an input for dementia and prostate cancer, breast cancer, MI and depression, so these scores are not methylation-only. The within-sex values for breast and prostate cancer are not from models retrained within sex, and may reflect reduced performance because of this. Finally, methylation-based scores for CVD have shown horizon dependence, with discrimination highest in the first years after sampling,^20^ and the ten-year horizon reported here should not be assumed to hold at longer follow-up.

Taken together, the results show that a single blood methylation profile can rank ten-year risk for several common diseases at a level that compares favourably with published genetic and methylation scores, and that it does so on the same assay that yields biological age, immune composition and surrogate biomarker measures. We report all 21 scores with confidence intervals and event counts so that the weak scores are visible alongside the strong. Further work is needed to expand the cohorts and training datasets, calibrate the scores for absolute risk, compare them directly with polygenic scores in the same participants, and test longitudinally whether they change with treatment, the property that could give them a place in care that genotype cannot match.

## METHODS

### Study populations

We developed methylation risk scores (MRS) for 21 diseases in 12,785 whole-blood DNA methylation samples measured on the Illumina Infinium MethylationEPIC v1 array from two cohorts. Generation Scotland (GS; n = 10,156)^28,29,69^ is a population-based cohort that recruited participants from 2006 - 2011 and sampled blood at the baseline clinic visit, with outcomes from linkage to national health records. Mass General Brigham (MGB; n = 2,629)^30^ is a hospital-based cohort that recruits participants through clinical care, and outcomes come from the electronic health record. Each individual contributed one sample. Mean age at collection was 49.3 years (SD 15.4) in GS and 57.2 (SD 16.0) in MGB; 56.1% of GS and 64.1% of MGB participants were female (Table 1).

Samples were split once into 70% training (n = 8,945), 15% validation (n = 1,920) and 15% holdout (n = 1,920) partitions; training and validation were pooled as the development set (n = 10,865) and the holdout was used only for final evaluation (see Data partitioning). The partitions were similar in composition: GS made up 79.5%, 79.3% and 79.4% of the training, validation and holdout partitions, mean age was 50.8-51.0 years and 57.7-57.8% were female. Control follow-up is the range across the 21 diseases, from observed follow-up only (Table 2).

#### DNA methylation preprocessing

Raw IDAT files were read with the SeSAMe Bioconductor package (v1.18+)^70^ and processed per sample with a custom pipeline. To detect color channel switching (CCS) probes, signal intensities were first dye-bias-adjusted with dyeBiasNL; this adjustment was used only to compute the in-band to out-of-band ratio and was not carried forward. Type I probes whose in-band to out-of-band signal ratio was below 1.5 were flagged as CCS probes. Second, a definitive non-linear dye-bias correction (dyeBiasNL) was fitted excluding the identified CCS probes and the SeSAMe-recommended EPICv1 masked probes, and then applied to all probes. Third, detection p-values were computed with pOOBAH, using out-of-band and negative-control signal as background and excluding CCS and recommended-masked probes from the background estimate. Probes with p-value > 0.05 were masked. Fourth, background subtraction was performed with noob^71^, with the same probe exclusions applied to the background estimate. Finally, beta values were extracted for all probes. CCS probes and probes with pOOBAH p-value > 0.05 were set to NA in the final beta matrix. EPICv1 probe identifiers are unique, so no replicate-probe collapsing was applied. Masked values were imputed with the probe median for feature ranking (see below).

### Sample quality control

Samples were excluded using a two-stage filter. In the first stage (QC1), samples were removed if total intensity was ≤ 5,000, detection success was ≤ 0.85, or the proportion of bad CpGs was ≥ 0.05. In the second stage (QC2), the distribution of beta values across ten bins of width 0.1 (0–0.1, …, 0.9–1.0) was computed for each sample. Samples were excluded if the proportion of probes in any bin fell outside the mean ± 4 SD of the samples that passed QC1. In total, 538 of 23,269 samples (2.31%) were excluded: 263 at QC1 and 275 at QC2.

#### Outcomes and follow-up

Twenty-one diseases were modelled (Table 2). Diseases were retained if more than 200 incident cases within 10 years of sample collection were available in the full GS and MGB pool. Disease definitions were harmonised across GS record-linkage codes and MGB electronic health record definitions (Supplementary Table 1). The modelling set (n = 12,785) comprised all participants with a diagnosis of at least one of the 21 diseases, plus the 2,233 GS participants flagged as healthy, who have no health record activity in the linked data. Participants with none of the 21 diagnoses and no healthy flag were not used, because cases limit class-balanced training and controls beyond four times the number of cases are subsampled in every training fold. Prevalent cases at sample collection were excluded from each disease-specific analysis. Controls were all remaining participants without a recorded diagnosis of the disease during follow-up, irrespective of other conditions; they were not restricted to disease-free individuals, so that the comparator matches the population in which a score would be applied. The time scale was years from sample collection to first diagnosis (cases) or to the end of follow-up (controls). Control follow-up differed between cohorts: median 15.1-15.2 years in GS (observed linkage follow-up) and 6.6-6.8 years in MGB (Table 1). Because the healthy-flagged participants have no linked records, they were assigned a censoring time of 12.0 years, just under the longest observed time to diagnosis among cases (about 13 years), which exceeds the 10-year evaluation horizon. For 24 MGB T2DM cases, event times were set to the recorded diagnosis dates. Outcomes were taken from linked health records (GS) and the electronic health record (MGB), recorded independently of and without knowledge of the methylation data, so outcome assessment was blind to the predictors.

#### Data partitioning

Samples were assigned once to training (n = 8,945), validation (n = 1,920) and holdout (n = 1,920) partitions (70/15/15) at the sample level, before any feature selection, so that the holdout could be used for an internal evaluation. Cohort mix, age and sex were balanced across the training, validation and holdout partitions. Training and validation samples were pooled as a development set (n = 10,865 before exclusion of prevalent cases) for feature selection and model fitting, with five-fold cross-validation within it. The holdout was used only to evaluate final models. Hyper-parameters of individual model members were tuned within cross-validation folds (see Survival models).

#### Probe selection

Within-array probe reproducibility was quantified using EPICv1 within-beadchip paired technical replicates (n = 29). Intraclass correlation coefficients (ICC) were computed using two-way random effects models with absolute agreement, ICC(2,1)^72^, with an inclusion threshold of ICC (2,1) > 0.7, implemented via the irr R package.^73^ Probes on chromosomes X and Y were also excluded. A variance filter retained probes with a standard deviation of beta values above 0.02, leaving 440,283 probes in the final pool.

#### Feature ranking and panel construction

Feature selection was based on comparing feature importances to null features using randomly instantiated tree-based classification models. Tree models were randomly instantiated 40 times, with a random 80/20 train-test split within each iteration. During each iteration, 20 random features were shuffled to remove any possible association with disease and create the null feature set. Feature importances for the null feature set and our real features were then computed using absolute TreeSHAP values on the test group.^74^ Features were retained only if they demonstrated a median TreeSHAP value >= 10 of the null feature sets median TreeSHAP values. Panels contained 119 (allergic rhinitis) to 962 (coronary artery disease) features, including age at collection where selected. Feature selection and the null threshold were pooled across cohorts.

#### Survival models

Models were fitted within the development set using five-fold cross-validation. In each fold, about 80% of the development samples formed the training fold and the remaining 20% the validation fold, which was used to tune model penalties. The holdout was not used at any stage of fitting or tuning.

Within each training fold, the classes were balanced: controls were undersampled to at most four times the number of cases, and cases were then oversampled with replacement to the number of controls (1:1). Validation folds and the holdout were not resampled. The scores therefore rank individuals by risk, but their absolute values are not population-calibrated.

For each disease, three survival models with a 10-year horizon were fitted: an elastic-net penalised Cox regression^75^, an ensemble of penalised Cox regressions (CoxPHEnsemble) and extra survival trees.^76^ The CoxPHEnsemble comprised 500 members (100 per fold). Each member was trained on a bootstrap resample of the balanced training fold, using a random subset of the disease’s panel of about one feature per 15 events (minimum five), with its penalty strength tuned on the validation fold. Predicted 10-year risks were averaged across members to give the risk score.

#### Evaluation

Each score was evaluated on the holdout at a 10-year horizon. Discrimination was measured with the inverse-probability-of-censoring-weighted concordance index (C-index)^77^ truncated at 10 years, which estimates the probability that the person diagnosed earlier was assigned the higher risk, and with the cumulative/dynamic area under the receiver operating characteristic curve (AUC) at 10 years, which compares individuals diagnosed within 10 years with those event-free at 10 years. Censoring distributions were estimated from the raw (unbalanced) training data. Ninety-five per cent confidence intervals were obtained from 10,000 bootstrap resamples of the holdout. For each disease, the reported model was the one of the three with the highest holdout C-index.

To present the results by strength of evidence (Table 3), scores were grouped into four descriptive tiers by the unrounded holdout C-index and the number of holdout events within 10 years: tier 1, C-index of at least 0.80 with at least 50 events; tier 1b, C-index of at least 0.80 with fewer than 50 events; tier 2, C-index from 0.70 to below 0.80; and tier 3, C-index below 0.70. The tiers describe point estimates and are not significance thresholds. All 21 scores are reported with confidence intervals and event counts irrespective of tier.

#### Hazard ratios

The 10-year risk score was standardised and a Cox proportional hazards model was fitted in the holdout over full follow-up, giving HRs per standard deviation of the score, adjusted for age and sex, and unadjusted (Table 3). Benjamini-Hochberg q-values were computed across fits. Because the score is a bounded probability, the unit is one standard deviation of that probability. For prostate and breast cancer, which occur in one sex, all metrics were calculated within the at-risk sex and hazard ratios were adjusted for age alone.

#### Feature attribution

SHAP values, which quantify how much each feature contributes to an individual’s predicted risk, were computed on the holdout for the CoxPHEnsemble of each disease. Because each ensemble member is a linear Cox model, its SHAP value for a feature is exact and equals the feature’s coefficient multiplied by the individual’s standardised deviation from the training mean. Values were averaged across the 500 members on the log-partial-hazard scale (zero where a member did not use the feature). As each feature is sampled by only a subset of members, features were ranked by the mean absolute SHAP value conditional on inclusion, that is, divided by the proportion of members that sampled the feature, among features sampled by at least 5 of 500 members. SHAP values show how much the model relies on a feature and do not indicate the direction or causality of an association. For dementia, the selected model was a Cox proportional hazards regression, for which SHAP values were computed in closed form and features with non-zero SHAP values in at least one cross-validation fold were retained. For CKD, the selected model was extra survival trees, for which interventional TreeSHAP values were computed on the 10-year predicted-probability scale, so their magnitudes are not comparable with those of other diseases.

#### Pathway and trait enrichment

Enrichment analyses were run on the panel of each of the 21 diseases, using the cg probes sampled by at least 5 of the 500 ensemble members, mapped to genes with the Illumina EPICv1 hg19 annotation. The background was the 439,530-probe variance-filtered pool from which the panels were selected (background probe panel of cg probes only). Over-representation of panel CpGs was tested in Gene Ontology biological process, KEGG, MSigDB Hallmark^78^, Reactome and WikiPathways gene sets with the missMethyl package^79^, which corrects for the number of probes per gene. P values were adjusted with the Benjamini-Hochberg method within each disease and gene-set collection (adjusted P < 0.05), without adjustment across diseases.^80^ Robustness was assessed by repeating the analysis on the top 25% and 50% of each panel by conditional SHAP value and with all EPICv1 probes as background. Panels were also compared with published whole-blood EWAS associations from the EWAS Atlas (hypergeometric test) and, as a supplementary analysis, tested by preranked gene set enrichment on the panel’s own genes. For the eight tier 1 and 1b diseases, panels were also compared with CpGs reported for the same disease in 21 published blood-based epigenome-wide association and methylation-risk-score studies (hypergeometric test). Panel CpGs were also compared with the CpGs that define 12 blood cell types in the EPIC reference library of the EpiDISH package^11,81^ (hypergeometric test against the same background). Genes were ranked by the largest conditional SHAP value among their panel CpGs, using the same hg19 annotation; CpGs without an hg19 gene were not ranked. Panel CpGs were classed as associated with age, sex or smoking if the CpG was reported for that trait in blood in the EWAS Atlas (P < 1e-7).^31^

#### Benchmarking against published scores

We compared the holdout performance of the 21 scores with published methylation risk scores (MRS) and polygenic risk scores (PRS) for the same diseases. A published score was eligible for inclusion if it was a multi-CpG methylation score or a multi-variant polygenic score for one of the 21 diseases and reported an AUC, C-index, odds ratio or hazard ratio for that disease. The following were excluded: Epigenetic clocks and other aging measures, standalone protein EpiScores and multi-omic scores, single-CpG or single-gene markers, epigenome-wide association studies without a score, cell-free DNA and non-blood tissue, and clinical-only models. Models that combined a multi-CpG methylation score with other predictors, such as clinical risk factors or a protein EpiScore, were retained, and the additional components are stated in Supplementary Tables 2 and 3.

Each eligible value was classified as closest, context or not comparable. A closest comparison required the same disease (or, for polygenic scores, its main subtype, for example idiopathic pulmonary fibrosis for interstitial lung disease and Alzheimer’s disease for dementia) and an AUC or C-index; for methylation scores it also required an incident design, with blood collected before diagnosis and prevalent cases excluded. For polygenic scores, prevalent, cross-sectional and case-control designs were accepted because genotype is fixed at birth. Context comparisons were informative but not like for like, for example a different endpoint or horizon, a composite endpoint, a population restricted to people with another disease, a preprint, a study that reported only odds or hazard ratios, or an estimate that the original authors described as circular, in-sample or possibly overfitted. Not comparable values were prevalent or cross-sectional methylation scores and non-blood or cell-free samples. Some published estimates may have been derived in the data used to select the score, CpGs, weights or tuning parameter (for example, a tuning parameter chosen by C-index in the evaluation cohort, or CpGs screened before a train-test split) and are therefore likely to be optimistic. Circularity was judged from the published methods, and overlap between evaluation and selection cohorts was assessed where reported. Where such an estimate otherwise met the closest criteria, it was retained as closest and marked as an exception, so that comparisons with our scores are conservative (ten of the 49 studies with a closest comparison; Supplementary Tables 2 and 3, and † in Table 4). Values from one study could fall into different classes.

Values were taken as published from the primary paper. The C-index was preferred and the AUC reported alongside it. Our scores include age and sex only where the model selected them, so published values that include covariates make the comparison conservative. Whether the training or validation cohort of a published score overlapped GS or the MGB was noted where reported. The difference between our value and the best closest published value was computed separately for methylation and polygenic scores, for score-alone and covariate-adjusted published values, using the same metric.

## SUPPLEMENTARY INFORMATION

All supplementary tables and figures are in a separate document.

## ETHICS STATEMENTS

This study was a secondary analysis of existing, de-identified data from two cohorts and involved no new participant recruitment, and was conducted in accordance with all applicable ethical guidelines and regulations. For MGB, ethical approval was obtained by the Mass General Brigham Institutional Review Board (protocol 2026P001153), and informed consent was obtained from all participants prior to their inclusion in the study. Data were anonymized to protect participant privacy, and confidentiality was strictly maintained throughout the study. All human subject research adhered to the principles outlined in the Declaration of Helsinki, ensuring respect for individuals, beneficence, and justice.

All components of Generation Scotland received ethical approval from the NHS Tayside Committee on Medical Research Ethics (REC Reference Number: 05/S1401/89). Generation Scotland has also been granted Research Tissue Bank status by the East of Scotland Research Ethics Service (REC Reference Number: 25/ES/0013), providing ethical approval for a wide range of uses within medical research. Written informed consent was obtained from all participants in GS:SFHS. This study was performed in accordance with the Helsinki declaration.

## DATA AVAILABILITY

The data from GS and the MGB Biobank cannot be shared publicly because of participant consent and data access agreements. According to the terms of consent for Generation Scotland participants, access to data must be reviewed by the Generation Scotland Access Committee. Access requests can be made through the researcher portal (https://gsaccess.igc.ed.ac.uk) and normally take up to six weeks for approval. Further information can be found on the Generation Scotland website: https://genscot.ed.ac.uk/for-researchers/access. MGB data can be shared for research purposes upon request via a Data Use Agreement by email request to Jessica Lasky-Su.

## AUTHOR CONTRIBUTIONS

R.S., M.Ma. and S.H. conceived the study. S.H., K.B.S., N.C.-G., L.B.-D. and V.D. developed the methodology and validated the work. S.H. built the models and performed the primary analysis, including the hazard ratios.

K.B.S. performed the additional analyses, including gene-set enrichment and benchmarking against published scores, and created the figures. R.E.M. and A.R. provided and curated the Generation Scotland data, and M.Me., A.A., T.G., K.L.I. and J.L.S. provided and curated the MGB data. K.B.S. and R.S. wrote the original draft. N.C.-G., L.B.-D., S.H., V.D., A.R., R.S., R.E.M., J.L.S. and K.L.I. reviewed and edited the manuscript. M.Ma. and R.S. supervised the work.

