## Supplementary Tables and Figures for "One blood methylation profile stratifies ten-year risk across 21 common diseases"

**Supplementary Table 1.** Disease definitions harmonised across Generation Scotland (GS) and the Mass General Brigham Biobank (MGB).

| Disease | Harmonised name | GS source name(s) | MGB source name | ICD codes? | Notes |
|---|---|---|---|---|---|
| **CKD** | Chronic_kidney_disease | ESRD, GN, obstr_reflux | Chronic_kidney_disease (native) | Yes | ESRD explicitly covered via ICD-10 N18. / ICD-9 585. |
| **PVD** | Peripheral_vascular_disease | peripheral_arterial_disease | Peripheral_vascular_disease (native) | Yes | GS code also feeds CVD_excl_stroke |
| **COPD** | COPD | COPD | COPD (native) | Yes | - |
| **CHF** | Congestive_heart_failure | hf | Congestive_heart_failure (native) | Yes | GS code also feeds CVD_excl_stroke |
| **T2DM** | Diabetes | diabetes (via Type2_Diabetes; GS has no Type 1 data) | Type1_Diabetes + Type2_Diabetes (native, unioned) | Partial (Type2_Diabetes only) | Derived composite (Type1 OR Type2), not a direct DISEASE_MAP entry. Byte-for-byte identical to Type2_Diabetes on every GS row, since GS's Type1_Diabetes is always 0. Only ~10 MGB patients (real Type 1 diagnoses) make the two columns diverge - treat as one signal, not two, for the GS-heavy portion of this cohort. |
| **Prostate cancer** | Prostate_cancer | pri_prost | Prostate_cancer (native) | No | - |
| **ILD** | Interstitial_lung_disease | pulm_fibrosis | Interstitial_lung_disease (native) | Yes | - |
| **Dementia (non-specific)** | NonSpec_Dementia | dementia | NonSpec_Dementia (native) | Yes | GS source (dementia) also maps to the removed CognitiveDeficit column - confirmed byte-for-byte identical across all 18,858 GS rows. Treated as one signal, not two. |
| **NAFLD / MASLD** | NAFLD | fatty_liver | NAFLD (native) | Yes | Also feeds Chronic_liver_disease for GS (100% deterministic subset) |
| **Breast cancer** | Breast_cancer | pri_breast | Breast_cancer (native) | No | - |
| **CAD** | Coronary_artery_disease | CHD_NOS, stable_angina, unstable_angina | Coronary_artery_disease (native) | Yes | Subset of CVD_excl_stroke's GS codes |
| **Lung cancer** | Lung_cancer | pri_lung | Lung_cancer (native) | No | - |
| **Stroke** | Stroke | Stroke_NOS, Isch_stroke, Intracereb_haem, Subarach | Stroke (native) | No | - |
| **MI** | Myocardial_infarction | myocardial_infarction | Myocardial_infarction (native) | No | GS code also feeds CVD_excl_stroke |
| **RA** | Rheumatoid_arthritis | RhA | Rheumatoid_arthritis (native) | Yes | - |
| **Depression** | Depression | depression | Depression (native) | Yes | - |
| **Allergic rhinitis** | Allergic_rhinitis | allergic_rhinitis | Allergic_rhinitis (native) | No | - |
| **Colorectal cancer** | Colon_Rectum_cancer | pri_bowel | Colon_Rectum_cancer (native) | No | - |
| **Asthma** | Asthma | asthma | Asthma (native) | Yes | - |
| **Psoriasis** | Psoriasis | psoriasis | Psoriasis (native) | No | - |
| **Alcohol-related liver disease** | Alcohol_liver_disease | liver_alc, alc_problems | Alcohol_liver_disease (native) | No | Only ~10% (GS) / ~43% (MGB) of cases also flag Chronic_liver_disease - most Alcohol_liver_disease cases come from alc_problems. |

*Abbreviations: GS, Generation Scotland; MGB, Mass General Brigham Biobank; ICD, International Classification of Diseases.*

*Each of the 21 modelled diseases was defined from the source disease fields of each cohort and harmonised to a common name. GS source names are the disease flags of the linked health records; MGB source names are the native electronic health record columns. "ICD codes?" indicates whether the definition is based on ICD codes. Further diseases available in MGB only (coin lesion of lung, hypertensive heart disease, chronic bronchitis, bronchiectasis, chronic pulmonary heart disease, chronic liver disease, and the composite cardiovascular disease excluding stroke) were not modelled and are not shown.*

### Supplementary Table 2. Our methylation risk scores (MRS) compared with published methylation scores.

| Disease | Published score [ref] | AUC | C-index | HR / OR per SD | Design | Comparability | Reason (context and not comparable) |
|---|---|---|---|---|---|---|---|
| **CKD** | Marchiori 2025 [1] | **0.82 (0.78–0.87)**, alone<br>**0.87 (0.83–0.92)**, + clinical | - | **1.10 (1.08–1.12)** per MRS unit, + clinical | Incident | Context | Participants with T2DM |
| | Jones 2024 [2] | **0.71**, alone<br>**0.92**, + clinical | - | **5.07 (2.48–10.38),** OR, + clinical<br>**1.66 (1.20–2.30)**, OR, + clinical | Cross-sectional | Not comparable | Prevalent, Cross-sectional |
| | Khurana 2026 [3] | **0.77 (0.71–0.82)**, alone<br>**0.78 (0.73–0.83)**, + clinical | - | - | Case-status | Not comparable | Different endpoint (Diabetic Kidney Disease) |
| | Li 2023 (DKD) [4] | **0.94**, + clinical<br>**0.88**, + clinical<br>**0.89**, alone<br>**0.81**, alone | - | - | Case-status / Cross-sectional | Not comparable | Prevalent, Cross-sectional, Different endpoint (Diabetic Kidney Disease) |
| | Thompson 2022 [5] | **0.898 (0.867–0.927)**, + covariates | - | - | Cross-sectional | Not comparable | Different endpoint |
| **PVD** | Barad 2026 (preprint) [6] | - | **0.736 / 0.743 / 0.734**, + covariates<br>**0.759 / 0.760 / 0.758**, + clinical | HR shown graphically<br>~**2.5–2.7** + covariates<br>~**2.0–2.1** + clinical | Incident | Context | Preprint. 89,6% males |
| | Ingold 2026 [7] | **0.92**, alone | **0.61**, + covariates | - | Prevalent / Incident | Not comparable | MACCE or prevalent PAD |
| **COPD** | Bermingham 2019 [8] | **0.856 (0.757–0.956)**, + clinical | - | - | Incident | Closest | - |
| | Wielscher 2015 [9] | **0.73 (0.62–0.83)**, alone | - | - | Case-control | Not comparable | cfDNA |
| **CHF** | Zhao 2022 [10] | **0.90 (0.88–0.92)**, + clinical<br>**0.65 (0.62–0.67)**, alone | - | - | Incident | Context | HFpEF endpoint |
| **T2DM** | Cardona 2019 [11] | **0.53**, alone<br>**0.762**, + clinical | - | - | Incident, apparent | Closest | CpGs selected in EPIC-Norfolk; weights possibly fitted in the evaluation sample, LOLIPOP (exception) |
| | Cheng 2023 [12] | **0.870**, + clinical<br>**0.872**, + clinical, epigenetic protein score | - | - | Incident, 10 y | Closest | - |
| | Fraszczyk 2022 [13] | **0.591**, alone<br>**0.757**, + clinical | - | - | Incident | Closest | - |
| | Huo 2024 [14] | **0.64**, alone<br>**0.635**, alone | - | **1.66 (1.29–2.14)** OR, + clinical | Incident | Closest | CpGs selected in the same sample (exception) |
| **Breast cancer** | Kresovich 2022 [15] | **0.63 (0.59–0.67)**, alone<br>**0.69 (0.63–0.75)**, alone<br>**0.71 (0.67–0.74)**, + clinical, PRS | - | **1.78 (1.51–2.11)**, + covariates | Incident | Closest | - |
| | Rodriguez 2026 [16] | **0.66 (0.61–0.72)**, alone<br>**0.71 (0.62–0.79)**, alone | - | - | Incident | Context | Different horizon, Selected on test data |
| | Wang 2022 [17] | **0.70**, + covariates | - | - | Case-status, apparent | Not comparable | Case-control, Prevalent |
| | Ruiz-De La Cruz 2024 [18] | **0.716**, alone<br>**0.929 (0.904–0.955)**, alone | - | - | Case-control | Not comparable | Case-control, hereditary-risk women, single CpG |
| **Prostate cancer** | Cheng 2026 (cfDNA) [19] | **0.73 (0.59–0.86)**, alone | **0.71**, alone | - | Incident | Not comparable | cfDNA |

| | | | | | | | |
|---|---|---|---|---|---|---|---|
| | Han 2021 [20] | **0.92**, alone | - | - | Prevalent | Not comparable | Different endpoint (aggressive vs non-aggressive prostate cancer, cases only) |
| | Moses-Fynn 2018 [21] | **0.87**, alone | - | - | Case-control | Not comparable | Prevalent, Case-control, single-gene |
| **ILD** | Wielscher 2015 [9] | **0.815 (0.73–0.88)**, alone | - | - | Case-control | Not comparable | cfDNA |
| **Dementia (non-specific)** | Hackenhaar 2023 [22] | **0.78**, + clinical<br>**0.99**, + clinical<br>**0.91**, + clinical<br>**0.772**, + covariates | - | - | Incident case-control, apparent / Cross-sectional | Context | Incident values (including the 0.78 pre-AD score) are in-sample (partly circular); the out-of-sample value (0.772, AIBL) is cross-sectional |
| | Koetsier 2024 [23] | **0.68 (0.60-0.77)**, alone | - | - | Cross-sectional | Not comparable | Prevalent MCI status |
| | Zhang 2025 [24] | - | - | **1.15 (1.006–1.32)** per MRS unit, + clinical | Incident | Context | No AUC / C-index to be the closest comparison. Different endpoint |
| | Acha 2023 [25] | **0.87**, + clinical | - | - | Case-control | Not comparable | Case-control, Prevalent |
| | Silva 2022 [26] | **0.696 (0.616–0.770)**, + covariates<br>**0.688**, + covariates<br>**0.614**, alone | - | - | Cross-sectional | Not comparable | Prevalent, Case-control |
| **NAFLD / MASLD** | Xia 2024 [27] | **0.77 (0.70–0.83)**, alone<br>**0.77 (0.58–0.96)**, + clinical, cross-validated<br>**0.81 (0.76–0.87)**, + clinical | - | - | Incident, apparent | Closest | CpGs selected in the same participants (exception) |
| **CAD** | Barad 2026 (preprint) [6] | - | **0.662 / 0.674 / 0.661**, + covariates<br>**0.694 / 0.699 / 0.694**, + clinical | HR shown graphically<br>**~1.7–1.9** + covariates<br>**~1.5–1.65** + clinical- | Incident | Context | Preprint. 89,6% males |
| | Cappozzo 2022 [28] | **0.721 (0.661–0.781)**, + clinical<br>**0.622 (0.551–0.693)**, alone | - | - | Incident, 10 y | Context | Composite CVD endpoint |
| | Westerman 2020 [29] | **0.642 (0.599–0.685)**, alone<br>**0.695 (0,655–0,734)**, + clinical | - | **1.28 (1.10–1.50)**, + covariates<br>**1.29 (1.09-1.51)** + covariates | Incident | Context | Composite CVD endpoint (coronary heart disease, stroke, CVD death) |
| | Chybowska 2024 [30] | **0.723**, + clinical | - | - | Incident, 10 y | Not comparable | Protein EpiScore, composite CVD |
| | García-Calzón 2025 [31] | **0.81 (0.77–0.86)**, alone<br>**0.84 (0.79–0.88)**, + clinical | - | - | Incident | Not comparable | T2D-only, composite endpoint |
| | Longo 2026 [32] | **0.936 (0.808–1.000)**, + covariates | - | - | Incident | Not comparable | PBMC, T2DM population |
| | Qin 2025 [33] | **0.71**, alone<br>**0.81**, + covariates<br>**0.83**, + clinical | - | - | Case-status | Not comparable | Different endpoint |
| | Zhang 2022 (CHD) [34] | **0.768 (0.694–0.843)**, alone<br>**0.834 (0.770–0.897)**, alone | - | - | Case-control | Not comparable | Case-control, Cross-sectional |
| **Lung cancer** | Bhardwaj 2026 [35] | **0.85 (0.80–0.90)**, alone | - | 4.88 (3.30-7.21) OR, alone | Incident | Closest | - |
| | Zhang 2016 (ESTHER) [36] | **0.80 (0.737–0.861)**, alone<br>**0.799 (0.733–0.866)**, alone<br>**0.789 (0.725–0.853)**, alone<br>**0.812 (0.725–0.871)**, alone | - | - | Incident | Closest | - |

| | | | | | | | |
|---|---|---|---|---|---|---|---|
| | Baglietto 2017 [37] | **0.817**, + clinical<br>**0.665**, + clinical | - | - | Incident | Closest | Selected on test data (exception) |
| | Fasanelli 2015 [38] | **0.78**, + clinical<br>**0.76**, + clinical | - | - | Incident | Closest | Selected on test data (exception) |
| | Onwuka 2024 [39] | **0.73 (0.68–0.77)**, alone | - | - | Incident | Closest | - |
| | Dawes 2026 [40] | **0.729**, + clinical | - | - | Incident | Context | Matched nested case-control, 20-yr horizon, single CpG |
| | Philibert 2022 [41] | **0.656 (0.600–0.709)**, + clinical | - | - | Incident | Context | Single CpG (cg05575921) with age and pack-years |
| | Wielscher 2015 [9] | **0.91 (0.84–0.96)**, alone | - | - | Case-control | Not comparable | Plasma cfDNA |
| | Zhao 2025 (lung) [42] | **0.55 (0.50–0.60)**, alone<br>**0.54 (0.50–0.59)**, alone<br>**0.57 (0.52–0.62)**, alone<br>**0.57 (0.49–0.65)**, alone<br>**0.55 (0.47–0.63)**, alone | - | - | Incident | Not comparable | Single CpG |
| **Stroke** | Barad 2026 (preprint) [6] | - | **0.719** / **0.721** / **0.721**, + covariates<br>**0.742** / **0.740** / **0.744**, + clinical | HR shown graphically<br>**~2.2–2.4** + covariates<br>**~1.8–1.9** + clinical- | Incident | Context | Acute ischemic stroke, 89,6% males |
| | Ingold 2026 [7] | - | **0.52**, + covariates | **1.03 (0.80–1.32)**, + covariates | Prevalent / Incident | Context | Peripheral atherosclerosis score |
| | Liu 2022 (HTRA1) [43] | **0.76 (0.67–0.84)**, + clinical<br>**0.66**, alone | - | - | Case-control | Not comparable | Different horizon, Small test set |
| | Sun 2022 [44] | **0.938**, + clinical | - | - | Case-control, apparent | Not comparable | Case-control, Prevalent |
| | Zhang 2022 (stroke) [45] | **0.84 (0.77–0.91)**, + clinical | - | - | Prognosis | Not comparable | Different endpoint (death after stroke) |
| **MI** | Ingold 2026 [7] | - | **0.63**, + covariates | **1.22 (1.02–1.47)**, + covariates | Prevalent / Incident | Context | Peripheral atherosclerosis score |
| | Long 2024 [46] | **0.686**, + clinical<br>**0.729**, + clinical | - | - | Incident | Context | Case-control, Different endpoint |
| **RA** | Pitaksalee 2023 [47] | **0.944**, + clinical<br>**0.93**, + clinical | - | - | Case-status | Not comparable | PBMC, apparent |
| | Prideaux 2025 (TIP-RA) [48] | **0.872**, alone | - | - | Incident | Not comparable | Preprint, Selected on test data |
| | Riskedal 2025 [49] | **0.96 (0.91–1.00)**, + clinical | - | - | Case-control | Not comparable | Case-control, Cross-sectional |
| | Thompson 2022 [5] | **0.626**, alone | - | - | Cross-sectional | Not comparable | Imputation / drug-response score |
| **Depression** | Barbu 2021 [50] | **0.58**, alone (prevalent)<br>**0.55**, alone (incident) | - | - | Prevalent / Incident | Closest | Only incident score is comparable |
| | Barbu 2022 [51] | **0.553**, alone<br>**0.569**, alone<br>**0.585**, alone | - | - | Prevalent | Not comparable | Prevalent, Cross-sectional |
| | Clark 2020 [52] | **0.724**, alone<br>**0.742**, + clinical | - | - | Incident | Not comparable | Different endpoint |

| | | | | | | | |
|---|---|---|---|---|---|---|---|
| | Shen 2025 [53] | **0.53**, alone | - | - | Case-status | Not comparable | EHR case-control |
| | Sokolov 2024 [54] | **0.569**, alone<br>**0.76**, alone | - | - | Case-status | Not comparable | Case-control, Cross-sectional; 0.76 from batch-level processed data |
| | Wang 2022 (mDI) [17] | **0.88**, alone<br>**0.60**, alone | - | - | Case-control, apparent / Case-control | Not comparable | Case-control, Prevalent |
| **Allergic rhinitis** | van Breugel 2022 [55] | **0.86**, alone | - | - | Prevalent | Not comparable | Nasal tissue |
| **Colorectal cancer** | Onwuka 2020 [56] | **0.82 (0.76–0.88)**, alone | - | - | Incident | Closest | CpGs possibly screened before the train-test split (exception) |
| | Jung 2024 [57] | **0.702 (0.583–0.822)**, alone | - | - | Incident, apparent | Closest | Six CpGs selected in the same participants (exception) |
| | Li 2026 (CRC) [58] | **0.88 (0.81–0.95)**, alone<br>**0.85 (0.74–0.94)**, alone<br>**0.82 (0.72–0.92)**, + clinical | - | - | Case-control, apparent / Case-control | Not comparable | Cross-sectional, Case-control |
| **Asthma** | Huang 2026 [59] | **0.736 (0.69–0.771)**, + covariates | - | - | Case-control | Not comparable | PBMC |
| | Li 2022 [60] | **1.00**, alone<br>**0.95**, alone | - | - | Case-control | Not comparable | Not whole blood |
| | Lin 2020 [61] | **0.87**, alone<br>**0.99**, alone<br>**0.97**, alone | - | - | Prevalent | Not comparable | PBMC |
| **Psoriasis** | Deng 2022 [62] | **0.976**, + covariates | - | - | Prevalent | Not comparable | Different endpoint, Prevalent |
| **Alcohol-related liver disease** | - | - | - | - | - | - | - |

*Abbreviations: AUC, area under the receiver operating characteristic curve; CI, confidence interval; cfDNA, cell-free DNA; HFpEF, heart failure with preserved ejection fraction; HR, hazard ratio; MACCE, major adverse cardiac and cerebrovascular events; MRS, methylation risk score; OR, odds ratio; PBMC, peripheral blood mononuclear cells; PCE, pooled cohort equations; PCs, principal components; PRS, polygenic risk score; SD, standard deviation; T2D, type 2 diabetes; UKB, UK Biobank.*

*Values are as reported, with 95% CI in parentheses where given. The text after each value gives the covariates in the published model at three levels: alone (no covariates); + covariates (demographic, technical or genetic covariates only, for example age, sex, principal components, genotyping array, batch, cell-type proportions, study or trial arm); and + clinical (any clinical or biological risk factor or clinical risk score, for example BMI, lipids, blood pressure, glucose or HbA1c, diabetes, smoking or pack-years, family history, the pooled cohort equations, genotype, or a lifestyle score). Where a model also includes another score (for example a protein EpiScore or a polygenic score), this is stated. Values separated by "/" are model variants or cohorts as stated. AUC is compared with AUC and C-index with C-index. Reference numbers [n] are those in the Supplementary References list.*

*Comparability (cell colour): Closest (green), same disease and an AUC or C-index and, for methylation scores, an incident design (for polygenic scores, prevalent, cross-sectional and case-control designs are accepted because genotype is fixed at birth); Context (yellow), relevant but the endpoint, design, horizon or population differs, or the report is a preprint; Not comparable (red), shown for completeness only. Rows are ordered closest, context, not comparable within each disease.*

*Estimates that may have been derived in the data used to select the score, CpGs, weights or tuning parameter are optimistic. Where they otherwise met the closest criteria, they were retained as closest and marked "(exception)", so that comparisons are conservative. Estimates that the authors themselves describe as circular or possibly overfitted were classed as context. In this table the exceptions are Cardona 2019, Xia 2024, Huo 2024, Baglietto 2017, Fasanelli 2015, Onwuka 2020, and Jung 2024; Hackenhaar 2023 and Rodriguez 2026 are context. Barad 2026 is a preprint and is classed as context. Barad 2026 values separated by "/" are the three model variants (MRS-1A, MRS-1B, MRS-2); Ingold 2026 MI and stroke values are for the peripheral atherosclerosis score, adjusted for age and sex.*

*"Apparent" denotes an estimate obtained in the same sample used to develop or select the score, without independent validation.*

#### Supplementary Table 3. Our methylation risk scores (MRS) compared with published polygenic risk scores.

| Disease | Published score [ref] | AUC | C-index | HR / OR per SD | Design | Comparability | Reason (context and not comparable) |
|---|---|---|---|---|---|---|---|
| **CKD** | Bakshi 2023 [63] | **0.58 (0.57–0.60)**, alone / **0.69 (0.67–0.70)**, + clinical (KDIGO)<br>**0.64 (0.63–0.66)**, alone / **0.73 (0.72–0.75)**, + clinical (eGFR < 60)<br>**0.65 (0.62–0.68)**, alone / **0.79 (0.77–0.81)**, + clinical (eGFR < 45) | - | **1.37 (1.31–1.43)** OR, + clinical<br>**1.75 (1.66–1.84)** OR, + clinical<br>**1.70 (1.52–1.90)** OR, + clinical | Cross-sectional | Closest | - |
| | Khan 2022 [64] | **0.81**, + clinical<br>**0.62**, alone | - | **1.46 (1.43–1.48)** OR, + clinical | Case-status | Closest | - |
| | Rodwell 2026 [65] | **0.765**, alone<br>**0.866 (0.860–0.871)**, + clinical<br>**0.66 (0.656–0.665)**, alone<br>**0.797 (0.794–0.800)**, + clinical | - | - | Cross-sectional / Case-status | Closest | - |
| | Liu 2025 [66] | - | - | **1.23 (1.21–1.24)**, + covariates | Incident | Context | HR only |
| | Tremblay 2021 [67] | **0.64**, + clinical | - | - | Incident | Context | T2D-only population |
| | Steinbrenner 2023 [68] | - | **0.84**, + clinical | **1.22 (1.11–1.34)**, + clinical | Incident | Context | Different endpoint (kidney failure); CKD-only population |
| **PVD** | Flores 2025 [69] | - | **0.761**, + covariates | **1.66 (1.61–1.71)**, + covariates | Incident | Closest | - |
| | Wang 2022 (PAD) [70] | **0.72 (0.71–0.72)**, + covariates<br>**0.804**, + clinical<br>**0.76 (0.75–0.77)**, + clinical | - | - | Prevalent | Closest | - |
| | Al Said 2025 [71] | **0.662**, + clinical | - | - | Incident | Context | Different endpoint, Different horizon |
| | Omiye 2024 [72] | **0.909 (0.856–0.961)**, + clinical<br>**0.818 (0.727–0.910)**, + clinical<br>**0.804 (0.709–0.898)**, + clinical | - | - | Case-control / Case-status | Not comparable | Case-control, Prevalent |
| **COPD** | Moll 2020 [73] | **0.67 (0.66–0.68)**, alone<br>**0.80 (0.79–0.81)**, + clinical | - | **1.81 (1.74–1.88)** OR, + clinical (European) | Case-control | Closest | - |
| | Liu 2025 [66] | - | - | **1.26 (1.24–1.28)**, + covariates | Incident | Context | HR only |
| **CHF** | Soh 2024 [74] | **0.561 (0.556–0.566)**, alone<br>**0.74 (0.736–0.744)**, + covariates | - | - | Incident | Closest | - |
| | Ahn 2025 [75] | - | **0.64 (0.626–0.655)**, + clinical<br>**0.714 (0.669–0.758)**, + clinical<br>**0.631 (0.617–0.644)**, + clinical | - | Incident | Context | Different endpoint, Different horizon |
| | Haller 2025 [76] | **0.798 (0.787–0.809)**, + covariates<br>**0.822 (0.811–0.832)**, + clinical<br>**0.829 (0.807–0.851)**, + clinical<br>**0.823 (0.798–0.848)**, + clinical | - | - | Incident | Context | Different horizon (3-year HF hospitalisation) |

| | | | | | | | |
|---|---|---|---|---|---|---|---|
| | | **0.822 (0.794–0.850)**, + clinical | | | | | |
| | Liu 2025 [66] | - | - | **1.49 (1.47–1.51)**, + covariates | Incident | Context | HR only |
| | Sinnott-Armstrong 2021 [77] | - | **0.635**, + covariates | **1.08 (1.06–1.10)**, + covariates | Incident | Context | Biomarker multi-PRS |
| | Tremblay 2021 [67] | **0.68**, + clinical | - | - | Incident | Context | T2D-only population |
| | Wang 2023 [78] | **0.75**, + covariates | - | - | Case-status | Context | Value from PGS Catalog, not reported in paper |
| **T2DM** | Drzymalla 2025 [79] | **0.739**, + clinical<br>**0.74**, + clinical<br>**0.785**, + clinical<br>**0.794**, + clinical | - | - | Incident / Case-status | Closest | - |
| | Huerta-Chagoya 2026 [80] | - | **0.85**, + clinical, European<br>**0.78**, + clinical, Admixed American | **1.28 (1.14–1.44)**, + clinical, African/African American<br>**1.71 (1.44–2.03)**, + clinical, Admixed American<br>**1.81 (1.70–1.94)**, + clinical, European | Incident | Closest | - |
| | Khera 2018 [81] | **0.73**, + covariates | - | - | Prevalent | Closest | - |
| | Mars 2020 [82] | - | **0.763 (0.758–0.767)**, + covariates<br>**0.845 (0.841–0.849)**, + clinical | - | Incident | Closest | Evaluated in FINRISK, the cohort used to choose the tuning parameter (exception) |
| | Thompson 2024 [83] | **0.694 (0.688–0.700)**, alone<br>**0.704 (0.698–0.710)**, alone | - | **2.131** OR, alone<br>**2.175** OR, alone | Case-status | Closest | - |
| | Wong 2022 [84] | **0.595 (0.575–0.615)**, alone<br>**0.638 (0.619–0.657)**, + covariates | - | - | Incident | Closest | - |
| | Hanson 2026 [85] | - | - | - | Incident | Context | Delta C-index only |
| | Liu 2025 [66] | - | - | **1.87 (1.85–1.89)**, + covariates | Incident | Context | HR only |
| **Breast cancer** | Mars 2020 [82] | - | **0.737 (0.733–0.741)**, + covariates<br>**0.750 (0.746–0.753)**, + clinical | - | Incident | Closest | Evaluated in FinnGen, which includes the FINRISK tuning cohort (exception) |
| | Mavaddat 2019 [86] | **0.63**, + covariates | - | **1.61 (1.57–1.65)** OR, + covariates | Incident | Closest | - |
| | Thompson 2024 [83] | **0.666**, alone<br>**0.67**, alone | - | **1.809** OR, alone<br>**1.837** OR, alone | Case-status | Closest | - |
| | Liu 2025 [66] | - | - | **2.16 (2.12–2.21)**, + covariates | Incident | Context | HR only |
| **Prostate cancer** | Conti 2021 [87] | **0.836 (0.832–0.840)**, + clinical<br>**0.833 (0.828–0.837)**, + covariates | - | - | Case-status | Closest | - |
| | Mars 2020 [82] | - | **0.857 (0.855–0.859)**, + covariates<br>**0.866 (0.863–0.868)**, + clinical | - | Incident | Closest | Evaluated in FinnGen, which includes the FINRISK tuning cohort (exception) |
| | Thompson 2024 [83] | **0.703**, alone<br>**0.708**, alone | - | **2.122** OR, alone<br>**2.157** OR, alone | Case-status | Closest | - |
| | Liu 2025 [66] | - | - | **2.09 (2.05–2.13)**, + covariates | Incident | Context | HR only |

| | | | | | | | |
|---|---|---|---|---|---|---|---|
| **ILD** | Duckworth 2026 [88] | **0.80 / 0.81**, alone (TOPMed)<br>**0.57 / 0.57**, alone (UKBB)<br>**0.82 / 0.83**, + covariates (TOPMed)<br>**0.74 / 0.74**, + covariates (UKBB) | - | - | Case-status | Closest | - |
| | Moll 2023 [89] | **0.70**, alone (PRS excluding MUC5B region)<br>**0.80**, + clinical<br>**0.81**, + clinical | - | - | Case-control | Closest | - |
| **Dementia (non-specific)** | Leonenko 2021 [90] | **0.698**, alone<br>**0.741**, + clinical | - | - | Case-control | Closest | - |
| | Mostafaei 2026 [91] | **0.691 (0.68–0.70)**, alone<br>**0.78 (0.77–0.80)**, + covariates (APOE locus excluded)<br>**0.83 (0.83–0.85)**, + covariates (APOE locus included)<br>**0.84 / 0.80 / 0.74**, + covariates (APOE-ε4 carriers) | - | - | Incident / Case-status | Closest | - |
| | Tanigawa 2022 [92] | **0.723 (0.678–0.769)**, alone, Alzheimer's<br>**0.869**, + covariates, Alzheimer's<br>**0.627 (0.596–0.658)**, alone, all-cause dementia<br>**0.816**, + covariates, all-cause dementia | - | - | Case-status | Closest | - |
| | Thompson 2024 [83] | **0.689 (0.658–0.719)**, alone<br>**0.697 (0.667–0.727)**, alone | - | **1.954** OR, alone<br>**1.976** OR, alone | Case-status | Closest | - |
| | Liu 2025 [66] | - | - | **1.96 (1.90–2.02)**, + covariates | Incident | Context | HR only |
| | Mukadam 2022 [93] | - | - | **1.73 (1.69–1.77)** OR, + covariates<br>**1.21 (1.18–1.24)** OR, + covariates | Incident | Context | No AUC in paper |
| | Zhang 2026 (dementia, preprint) [94] | - | **0.816**, + covariates<br>**0.846**, + clinical | - | Incident | Context | Preprint |
| **NAFLD / MASLD** | Namjou 2019 [95] | **0.596**, alone | - | - | Case-control | Closest | - |
| | Xiao 2024 [96] | **0.581 (0.577–0.584)**, alone | - | - | Case-status | Context | Proxy-defined MASLD |
| | Schwantes-An 2024 [97] | **0.650 (0.619–0.680)**, + clinical | - | - | Case-control | Not comparable | Case-control, Prevalent |
| **CAD** | Elliott 2020 [98] | - | **0.61 (0.60–0.62)**, alone<br>**0.78 (0.77–0.79)**, + clinical | **1.32 (1.30–1.34)**, alone | Incident | Closest | - |
| | Inouye 2018 [99] | - | **0.623 (0.615–0.631)**, + covariates | **1.58 (1.55–1.61)**, + covariates | Incident | Closest | - |
| | Khera 2018 [81] | **0.81**, + covariates | - | - | Prevalent | Closest | - |
| | Mars 2020 [82] | - | **0.832**, + covariates,<br>**0.820**, + clinical | - | Incident, tuning set | Closest | Tuning-set value (exception) |
| | Riveros-Mckay 2021 [100] | - | **0.63**, alone | **1.62 (1.57–1.67)**, alone | Incident | Closest | - |
| | Patel 2023 [101] | - | **0.763 (0.759–0.768)**, + clinical | **1.73 (1.70–1.76)** HR, + covariates<br>**2.14 (2.10–2.19)** OR, + covariates | Incident / Prevalent | Closest | - |
| | Thompson 2024 [83] | **0.643 (0.636–0.651)**, alone<br>**0.665 (0.658–0.672)**, alone | - | **1.731** OR, alone<br>**1.876** OR, alone | Case-status | Closest | - |

| | | | | | | | |
|---|---|---|---|---|---|---|---|
| | Wong 2022 [84] | **0.587 (0.576–0.599)**, alone<br>**0.706 (0.696–0.716)**, + covariates | - | - | Incident | Closest | - |
| | Liu 2025 [66] | - | - | **1.46 (1.44–1.47)**, + covariates | Incident | Context | HR only |
| **Lung cancer** | Galal 2026 [102] | - | **0.569 (0.559–0.578)**, alone | **1.28 (1.23–1.32)**, alone | Incident | Closest | - |
| | Zhang 2026 (lung) [103] | **0.562**, alone<br>**0.563**, alone<br>**0.624**, alone<br>**0.589**, alone<br>**0.638**, alone<br>**0.595**, alone | - | - | Prevalent / Case-status / Case-control | Closest | - |
| | Blechter 2025 (JTO) [104] | **0.63**, alone | - | - | Case-control | Context | Case-control, Different endpoint |
| | Blechter 2026 (JNCI) [105] | **0.640 (0.629–0.653)**, alone<br>**0.629 (0.618–0.641)**, alone<br>**0.649 (0.623–0.676)**, alone<br>**0.653 (0.631–0.675)**, alone | - | **1.71 (1.61–1.82)** OR, alone | Case-control | Context | Case-control, Different endpoint |
| | Boumtje 2026 [106] | **0.629**, + covariates<br>**0.575**, + covariates<br>**0.654**, + covariates<br>**0.707**, + clinical | - | - | Case-status | Context | Screening cohort |
| | Liu 2025 [66] | - | - | **1.29 (1.25–1.33)**, + covariates | Incident | Context | HR only |
| | Lung PRS validation 2023 [107] | **0.588**, alone<br>**0.738**, + clinical | - | - | Case-control | Context | Case-control |
| | Ma 2025 [108] | - | **0.58**, + clinical | **1.33 (1.27–1.39)**, + covariates | Incident | Context | Abstract only |
| **Stroke** | Abraham 2019 [109] | - | **0.58 (0.566–0.593)**, alone<br>**0.637**, + clinical | - | Incident | Closest | - |
| | Rutten-Jacobs 2018 [110] | - | **0.69**, + covariates | - | Incident, apparent | Closest | Best of three SNP thresholds chosen in the same cohort (exception) |
| | Thompson 2024 [83] | **0.581 (0.566–0.596)**, alone<br>**0.596 (0.581–0.611)**, alone | - | - | Case-status | Closest | - |
| | Wong 2022 [84] | **0.512 (0.492–0.533)**, alone<br>**0.668 (0.650–0.686)**, + covariates | - | - | Incident | Closest | - |
| | Liu 2025 [66] | - | - | **1.49 (1.46–1.52)**, + covariates | Incident | Context | HR only |
| | Neumann 2021 [111] | **0.685 (0.640–0.730)**, + clinical | - | **1.41 (1.20–1.65)**, + clinical | Incident | Context | Different horizon |
| | Tremblay 2021 [67] | **0.66**, + clinical<br>**0.62**, + clinical | - | - | Incident | Context | T2D-only population |
| **MI** | Gunn 2025 [112] | - | **0.783**, + covariates | **1.54 (1.46–1.63)** OR, + covariates | Prevalent | Context | C-index and OR from PGS Catalog, not reported in paper |
| | Liu 2025 [66] | - | - | **1.34 (1.32–1.36)**, + covariates | Incident | Context | HR only |
| | Tremblay 2021 [67] | **0.67**, + clinical<br>**0.61**, + clinical | - | - | Incident | Context | T2D-only population |

| | | | | | | | |
|---|---|---|---|---|---|---|---|
| **RA** | Ishigaki 2022 [113] | **0.66**, + covariates | - | - | Case-control | Closest | - |
| | Tanigawa 2022 [92] | **0.665**, + covariates<br>**0.600 (0.584–0.616)**, alone | - | - | Case-status | Closest | - |
| | Thompson 2024 [83] | **0.599 (0.587–0.612)**, alone<br>**0.606 (0.593–0.619)**, alone | - | - | Case-status | Closest | - |
| | Jermy 2024 [114] | - | - | **1.64 (1.62–1.67)**, + covariates | Incident | Context | Different horizon |
| **Depression** | PGC MDD 2025 [115] | **0.625**, + covariates | - | - | Case-control | Closest | - |
| | Halldorsdottir 2019 [116] | **0.597**, alone<br>**0.631**, + clinical | - | - | Incident | Context | Different horizon |
| | Jermy 2024 [114] | - | - | **1.20 (1.19–1.20)**, + covariates | Incident | Context | Different horizon |
| | Liu 2025 [66] | - | - | **1.27 (1.25–1.28)**, + covariates | Incident | Context | HR only |
| | Mitchell 2021 [117] | - | - | **1.75 (1.73–1.77)** OR, + covariates | Case-control | Context | Case-control, Different endpoint |
| | Yu 2025 [118] | **0.63 (0.61–0.64)**, + clinical | - | - | Cross-sectional | Not comparable | Cross-sectional, Different endpoint |
| **Allergic rhinitis** | Leskien 2026 [119] | **0.64**, + clinical | - | - | Incident | Not comparable | Different endpoint, Different horizon |
| **Colorectal cancer** | Thomas 2020 [120] | **0.654 (0.639–0.669)**, + covariates | - | - | Case-control | Closest | - |
| | Thompson 2024 [83] | **0.631 (0.617–0.645)**, alone<br>**0.639 (0.625–0.652)**, alone | - | - | Case-status | Closest | - |
| | Liu 2025 [66] | - | - | **3.32 (3.24–3.41)**, + covariates<br>**3.09 (3.01–3.18)**, + covariates | Incident | Context | HR only |
| **Asthma** | Dapas 2023 [121] | **0.623**, alone | - | - | Prevalent | Closest | Best of five PRS chosen in the same population (exception) |
| | Sordillo 2021 [122] | **0.55**, alone<br>**0.54**, alone | - | - | Case-control | Closest | - |
| | Thompson 2024 [83] | **0.615 (0.610–0.620)**, alone<br>**0.642 (0.637–0.647)**, alone | - | - | Case-status | Closest | - |
| | Liu 2025 [66] | - | - | **1.13 (1.11–1.16)**, + covariates | Incident | Context | HR only |
| | Namjou 2022 [123] | **0.70 (0.69–0.72)**, + covariates<br>**0.66 (0.65–0.66)**, + covariates | - | - | Case-control | Context | Different endpoint |
| **Psoriasis** | Tanigawa 2022 [92] | **0.698**, + covariates<br>**0.692 (0.675–0.708)**, alone | - | - | Case-status | Closest | - |
| | Thompson 2024 [83] | **0.685 (0.672–0.698)**, alone<br>**0.702 (0.689–0.715)**, alone | - | - | Case-status | Closest | - |
| **Alcohol-related liver disease** | Ding 2023 [124] | - | - | - | Incident | Context | HR only |
| | Schwantes-An 2024 [97] | **0.679 (0.655–0.703)**, + covariates<br>**0.619 (0.592–0.646)**, + clinical<br>**0.664 (0.638–0.691)**, + clinical<br>**0.734 (0.685–0.783)**, + clinical | - | - | Case-control | Context | Case-control, Prevalent |

| | | | | | | | |
|---|---|---|---|---|---|---|---|
| | Sinnott-Armstrong 2021 [77] | - | **0.626**, + covariates | **1.18 (1.11–1.27)**, + covariates | Incident | Context | Biomarker multi-PRS |
| | Whitfield 2022 [125] | **0.619**, alone<br>**0.665**, alone | - | - | Case-status | Context | Case-control, Cross-sectional |

*Abbreviations: AUC, area under the receiver operating characteristic curve; CI, confidence interval; cfDNA, cell-free DNA; HFpEF, heart failure with preserved ejection fraction; HR, hazard ratio; MACCE, major adverse cardiac and cerebrovascular events; MRS, methylation risk score; OR, odds ratio; PBMC, peripheral blood mononuclear cells; PCE, pooled cohort equations; PCs, principal components; PRS, polygenic risk score; SD, standard deviation; T2D, type 2 diabetes; UKB, UK Biobank.*

*Values are as reported, with 95% CI in parentheses where given. The text after each value gives the covariates in the published model at three levels: alone (no covariates); + covariates (demographic, technical or genetic covariates only, for example age, sex, principal components, genotyping array, batch, cell-type proportions, study or trial arm); and + clinical (any clinical or biological risk factor or clinical risk score, for example BMI, lipids, blood pressure, glucose or HbA1c, diabetes, smoking or pack-years, family history, the pooled cohort equations, genotype or a lifestyle score). Where a model also includes another score (for example a protein EpiScore or a polygenic score), this is stated. Values separated by "/" are model variants or cohorts as stated. AUC is compared with AUC and C-index with C-index. Reference numbers [n] are those in the Supplementary References list.*

*Comparability (cell colour): Closest (green), same disease and an AUC or C-index and, for methylation scores, an incident design (for polygenic scores, prevalent, cross-sectional and case-control designs are accepted because genotype is fixed at birth); Context (yellow), relevant but the endpoint, design, horizon or population differs, or the report is a preprint; Not comparable (red), shown for completeness only. Rows are ordered closest, context, not comparable within each disease.*

*Many published PRS values were estimated as case-control or mixed prevalent and incident discrimination (for example UKB test subgroups), not as 10-year incident prediction; lifetime genetic risk is not time-limited. Estimates that may have been derived in the data used to select the score, CpGs, weights or tuning parameter are optimistic. Where they otherwise met the closest criteria, they were retained as closest and marked "(exception)", so that comparisons are conservative. Estimates that the authors themselves describe as circular or possibly overfitted were classed as context. In this table the exceptions are Mars 2020, Dapas 2023 and Rutten-Jacobs 2018.*

*Mars 2020: C-indices are from Table 1 of the paper (Cox models, incident cases, 10-year horizon): age + sex + PRS (age + PRS for breast and prostate cancer) and clinical risk score + PRS. CHD and T2D were evaluated in FINRISK and breast and prostate cancer in FinnGen, which includes the FINRISK participants. The LDpred tuning parameter was chosen by C-index in FINRISK (Supplementary Table S11), so the estimates are optimistic. Dapas 2023: the PRS with the highest AUC was selected in the same population.*

*Tanigawa 2022: values are from the hold-out test set (white British), Supplementary Table S6 of that paper; covariates are age, sex, genotyping array and the top ten genotype PCs. Alzheimer's disease and all-cause dementia are separate traits. Thompson 2024, Leonenko 2021 and Liu 2025 report Alzheimer's disease for dementia and ischaemic stroke for stroke.*

*Duckworth 2026: TOPMed and UKBB are replication cohorts; the Columbia discovery cohort and models that include MUC5B or rare variants are not shown. Moll 2023: values are for idiopathic pulmonary fibrosis. Namjou 2019: a 10-SNP score in the eMERGE cohort, which includes the Partners/Brigham site. Huerta-Chagoya 2026: the clinical score alone gave a C-index of 0.83 in European ancestry.*

*"Apparent" denotes an estimate obtained in the same sample used to develop or select the score, without independent validation.*

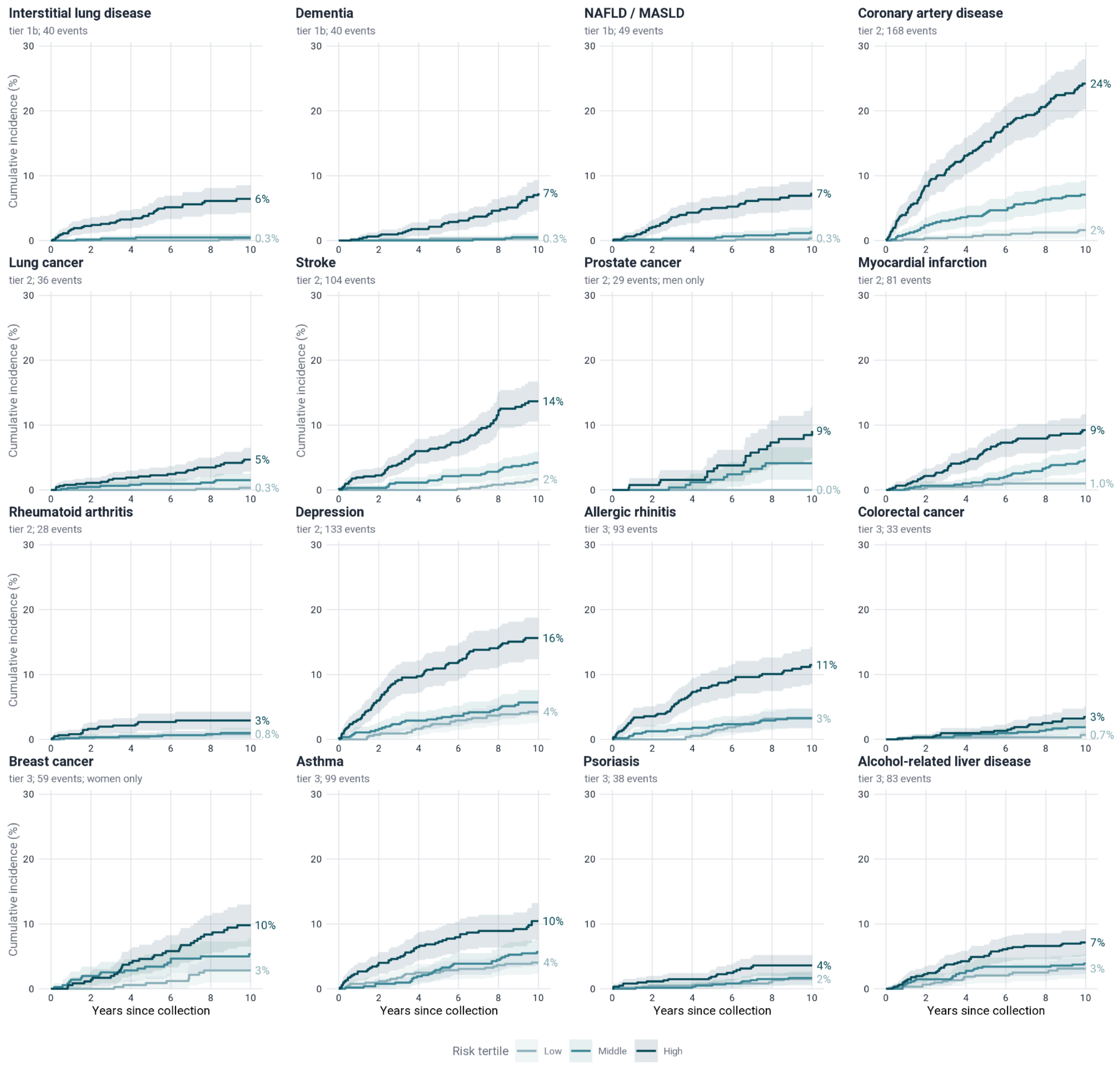


**Supplementary Figure 1.** Kaplan-Meier cumulative incidence over 10 years from sample collection, by tertile of the 10-year risk score in the holdout, for tier 1b (interstitial lung disease, dementia, NAFLD / MASLD), tier 2 (prostate cancer, coronary artery disease, lung cancer, stroke, myocardial infarction, rheumatoid arthritis, depression) and tier 3 (breast cancer, allergic rhinitis, colorectal cancer, asthma, psoriasis, alcohol-related liver disease) diseases. Methods are as in Fig. 2C: equal-sized tertiles with cutpoints computed within the holdout; follow-up truncated at 10 years; participants without the event censored at last follow-up or 10 years. Each panel gives the tier and the number of holdout events within 10 years; the y axis is shared. Shaded bands are pointwise 95% confidence intervals; values at the right are the 10-year cumulative incidence in the low and high tertiles. Prostate and breast cancer occur in one sex, so their curves are for men (n = 785) and women (n = 1,067) only, with tertiles cut within that sex. Several diseases have few events in the low tertile (none for prostate cancer), so those curves are imprecise. Scores are not calibrated, tertiles are relative to this cohort, and curves are not adjusted for age or sex.

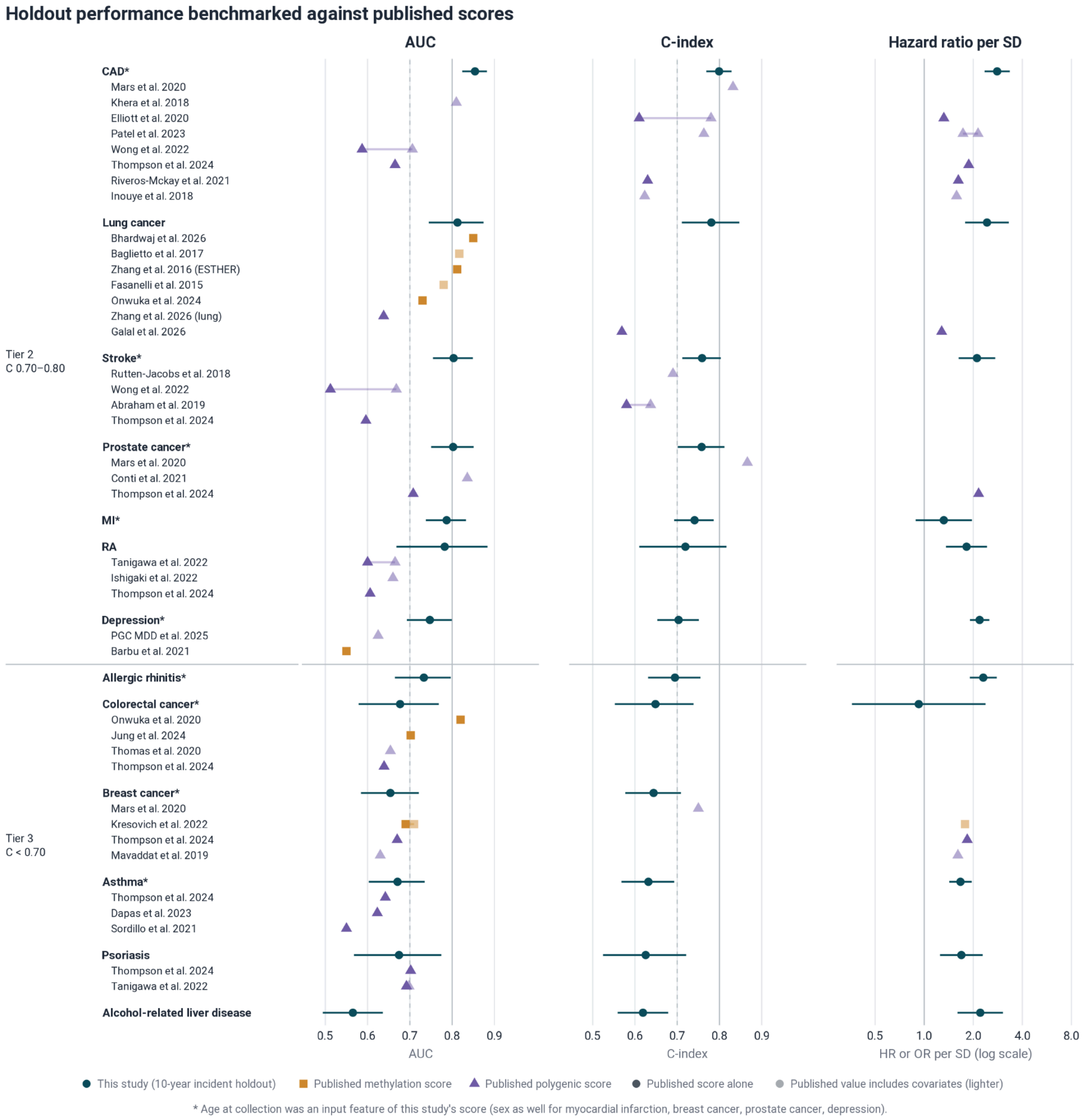


**Supplementary Figure 2.** Benchmark of the methylation risk scores against published methylation and polygenic risk scores for tier 2 and tier 3. As Fig. 3A for the remaining 13 diseases: tier 2 (coronary artery disease (CAD), lung cancer, stroke, prostate cancer, myocardial infarction (MI), rheumatoid arthritis (RA), depression) and tier 3 (allergic rhinitis, colorectal cancer, breast cancer, asthma, psoriasis, alcohol-related liver disease), with the same symbols, closest-comparison rule and axes. Diseases with no closest published score show this study only. * Panel includes age at collection as a covariate.
